# Predicting cardiovascular risk under intervention: Development and internal validation of the CHARIOT Model in 19 million adults

**DOI:** 10.1101/2025.09.18.25336064

**Authors:** Alexander Pate, Bowen Jiang, Yun-Ting Huang, Sophie Griffiths, David Stables, Niels Peek, Brian McMillan, Matthew Sperrin

**Author notes:** **<u>Corresponding author:</u>** Alexander Pate.

## Abstract

Cardiovascular disease (CVD) risk prediction models are widely used to guide primary prevention, yet conventional approaches can only tell patients that *something* should change, not how much specific actions would reduce their risk. Developed using electronic health records from 19,410,403 UK adults, CHARIOT combines survival analysis with causal inference to predict 10-year CVD risk under statin initiation, antihypertensive therapy, smoking cessation, or lifestyle modifications targeting weight, blood pressure and lipids, and is designed for repeated use over clinical encounters.

To illustrate its clinical utility: for a 70-year-old woman with 18.97% baseline risk, CHARIOT estimates risk reductions to 14.32% (statins), 15.26% (10mmHg blood pressure reduction), or 15.77% (smoking cessation). Internal validation demonstrated strong calibration across sex, age, ethnicity, and English regions, with discrimination (c-statistic) of 0.874 (female) and 0.859 (male). CHARIOT is publicly available as an interactive application and represents a substantive step toward actionable, patient-centred CVD prevention at scale.

## 1. Introduction

Cardiovascular disease (CVD) is the leading cause of death worldwide, and causes a quarter (>170,000) of all deaths in the UK every year, and costs the NHS and the UK economy an estimated £12 billion and £28 billion per year respectively.^1^ Models have been developed to predict incident CVD in various locations: QRISK^2^ (UK); ASSIGN^3^ (Scotland); SCORE2 and SCORE2-OP^4^ (Europe), Pooled Cohort Equations^5,6^ (United States); PREDICT-CVD^7^ (New Zealand), Globorisk^8^ (worldwide). These models estimate cardiovascular risk so that high risk individuals can be identified for intervention, often statins, and several are recommended in guidelines for primary prevention of CVD.^9–11^. In England, CVD risk is evaluated as part of the NHS Health Check, which is offered to individuals between ages 40 and 74 years that meet certain criteria.^12^ CVD risk is calculated using a prognostic model (QRISK), and those with a 10-year CVD risk of 10% or more are recommended to be offered a cholesterol lowering medication (statin) and given lifestyle advice on reducing their risk.^11,13^ The goal of this activity is prevention: to reduce the number of individuals developing CVD (and other chronic conditions), and to spur people on to make healthy lifestyle changes.^14^

Despite their widespread use, conventional CVD risk prediction models have a fundamental limitation as decision support tools: they are purely associational and therefore cannot answer the questions that matter most to patients and clinicians. When a patient asks “what would happen to my risk if I stopped smoking?” or “how much would statins actually help me?”, existing models cannot respond. Some clinical systems approximate an answer by applying published average treatment effects to an individual’s observed risk score,^15^ but this approach has well-recognised problems. Standard models do not appropriately handle treatment use in the development cohort,^16,17^ either ignoring it (resulting in risks that conflate treated and untreated individuals), or excluding individuals on treatment at baseline (e.g. statins), which prevents models from being applied at follow-up visits when treatment is already underway. Moreover, this approach cannot be extended to lifestyle interventions such as weight loss or dietary change. The result is a tool that can only tell patients and clinicians that *something* should change, but not what.

This matters because the way risk information is communicated influences whether patients change behaviour. People are more likely to change their behaviour when they perceive a personal risk and believe that specific actions will reduce it – a psychological mechanism known as response efficacy.^18^ Yet current tools, including QRISK and the Pooled Cohort Equations, provide no individualised information about the benefit of specific interventions.

Recent methodological advances combining risk prediction with causal inference, sometimes called “prediction under intervention,”^19^ now make it possible to estimate an individual’s risk under hypothetical interventions in a rigorous way, preserving the calibration and discrimination properties expected of a clinical prediction model while adding genuine decision support functionality. We developed CHARIOT to realise this potential in a large, representative English population.

Our aim was to develop a clinical prediction model that allows *prediction of CVD risk under a range of interventions* such as initiating statin or antihypertensive therapy, smoking cessation, changes in lifestyle such as diet or exercise, or ‘doing nothing’. This model should perform well in the classical clinical prediction sense (i.e. is well calibrated and discriminates well), whilst also having the functionality to ‘predict under intervention’. The intended target population is English adults free from CVD. The first intended setting for model use is when a patient meets with a general practitioner in primary care, for example during an NHS health check (although not restricted to this setting). The second intended setting for the model is through patient-facing online health records platforms.^20^ The goal of the model is to provide better information to support individuals and clinicians to make informed choices. If successful, this could help motivate behaviour change in order to prevent incident CVD events.

## 2. Methods

This study follows the TRIPOD+AI reporting guidelines,^21^see supplementary data file 3.

### 2.1. Overview of CHARIOT architecture

The CHARIOT model is designed to be used across multiple visits or interactions with the health system. It has two components: an initial risk-estimation component, and an intervention component. The initial-risk estimation component is a prediction model developed on a large linked primary care dataset, and is used to estimate an individual’s risk at their first visit. The intervention component adjusts predicted risk depending on interventions. It outputs both: actual risk predictions at subsequent visits, based on changes in risk factors since first visit; and hypothetical risks based on interventions under consideration to support decision-making. The intervention component is informed by effect estimates from a combination of trials and observational studies identified from the literature.

An example of how the CHARIOT model is used across the first two visits is given in Figure 1, extended to N visits in supplementary data file 1. This process is illustrated in a worked example in section 3.3.

**FIGURE 1:**
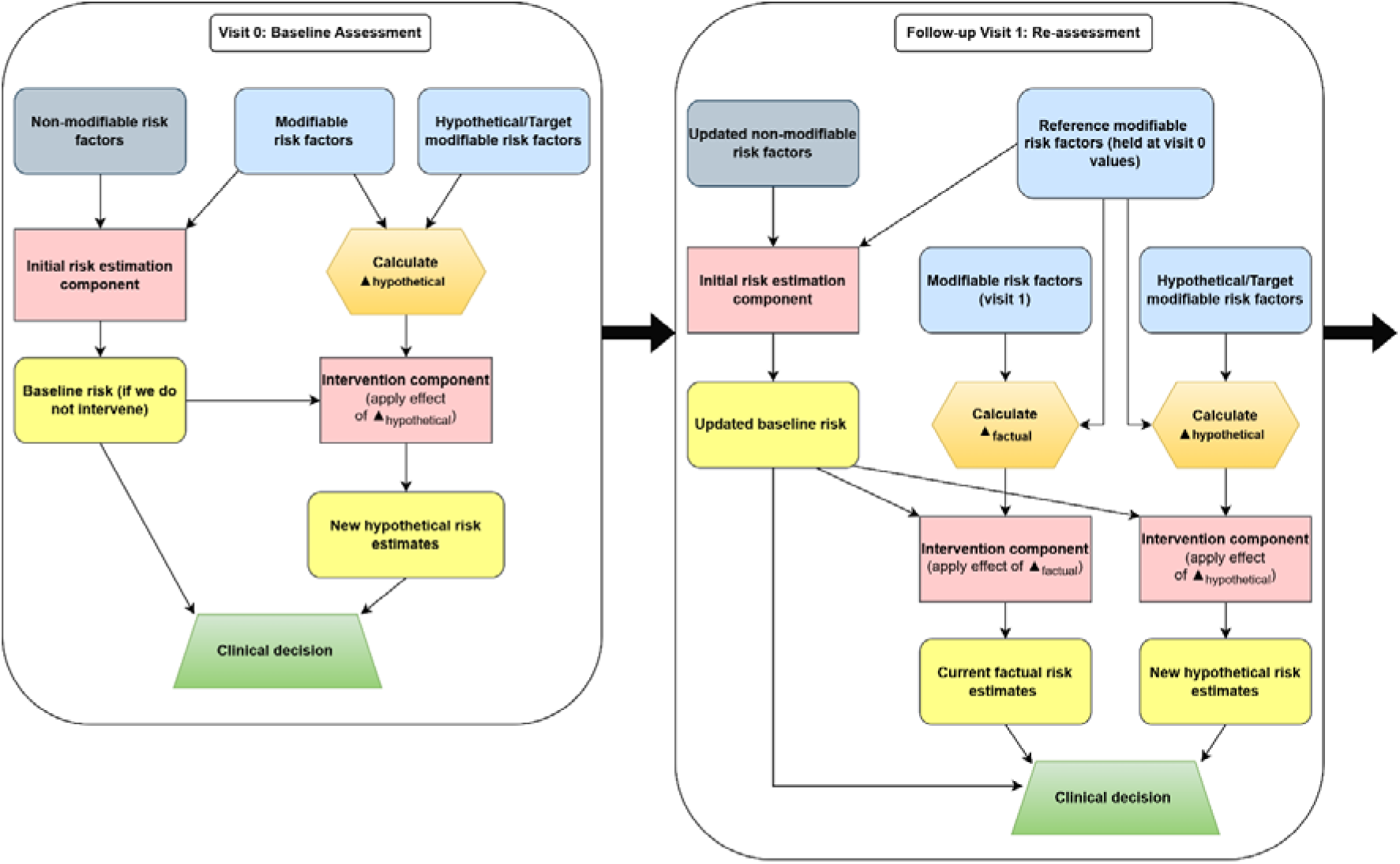
CHARIOT architecture across multiple visits.

### 2.2. Assumptions required for prediction under intervention

We make two key assumptions. We first assume the assumptions for estimation of causal effects (consistency, exchangeability and positivity) are met in the estimation of the causal effects borrowed from literature. Secondly, we assume transportability of the estimates borrowed from the literature to this population. These are both strong assumptions, and this is considered further in the discussion.

### 2.3. Development of the initial risk estimation component

#### 2.3.1. Study design

We conduct a retrospective cohort study using primary care data from Clinical Practice Research Datalink (CPRD) Aurum June 2021 extract. CPRD is a large resource of electronic health records (EHRs) from the UK containing information on demography, medical history, test results and drug use of individuals registered with a general practice. CPRD Aurum contains data from general practices using the EMIS Web computer system. As of June 2024, CPRD Aurum contained data on 47 million (16 million currently registered) individuals.^22^ Linkage was provided to secondary care data (Hospital Episode Statistics), death data (Office for National Statistics) and index of multiple deprivation.

This study was designed to match the proposed target population for primary prevention of CVD – of all CVD free adults in the England. The cohort comprised all individuals aged 18 or over registered during the study period, who had not yet been diagnosed with CVD. Individuals entered the cohort at the latest of: the start of the study period (1/1/2005), attaining at least one year of registration with a contributing practice to CPRD Aurum, attaining age 18 (referred to as ‘baseline index date’ or ‘visit 0’). Individuals exited the cohort at the earliest of: the date of diagnosis of first CVD event, death, deregistration with practice, last data upload from practice, or end of follow-up in HES (01/03/2020). Individuals were excluded if their cohort exit date was smaller than or equal to their cohort entry date, or they had a history of intracerebral haemorrhage prior to their cohort entry date. Note, we do not exclude individuals who have a record of statin use prior to their baseline index date. After applying all exclusion criteria, this resulted in individuals from 1478 English general practices. Access to all data was approved via CPRD’s Research Data Governance process (protocol ID 22_002333).

#### 2.3.2. Outcome

The outcome was incident CVD, where CVD was defined to be a composite outcome of coronary heart disease, ischaemic stroke or transient ischaemic attack, including death caused by any of these events. CVD events were identified through the primary care, secondary care, and office for national statistics death data. CVD events as a consequence of developing a different condition in hospital were not considered. Code lists and motivation for the definition of the outcome and the exclusion criteria (which includes intracerebral haemorrhage) are given in the supplementary data file 1.

#### 2.3.3. Exposures

Exposures were split into two groups: (i) modifiable risk factors and (ii) interventions. Modifiable risk factors were systolic blood pressure, body mass index, non-high-density-lipoproteins (non-HDL) cholesterol and smoking status. We considered any interventions which we expected to act through modification of these risk factors, e.g. smoking cessation, statin use, antihypertensive use and changes to diet and exercise. For statins and antihypertensives, an individual was considered to be “off treatment” 180 days after their last prescription, if they did not receive another prescription in that 180-day time period.

#### 2.3.4. Baseline predictors

Baseline predictors were variables used for estimation of the risk prior to intervention. The same set of predictor variables (with some minor differences) that were used in QRisk4^2^ were selected: age, sex, ethnicity, index of multiple deprivation (vigintile), systolic blood pressure, body mass index, smoking status, history of hypertension, atrial fibrillation, chronic kidney disease stage 3, 4 or 5, diabetes, serious mental illness, family history of CVD, migraine, systemic lupus erythematosus, corticosteroid use, atypical antipsychotic use, chronic obstructive pulmonary disease, intellectual disability, downs syndrome, oral cancer, brain cancer, lung cancer, blood cancer, pre-eclampsia (females only), postnatal depression (females only) and impotence (males only). These variables have been shown to result in a strong prediction model for CVD in a similar population and therefore no variable selection was applied. Systolic blood pressure variability was not considered given the role of systolic blood pressure as a modifiable risk factor which will be treated causally (see Section 2.3). Non-HDL cholesterol was included rather than cholesterol/HDL ratio due to the availability of causal estimates for changes in this modifiable risk factor (see supplementary data file 1). We also included a calendar time variable given the secular trend in CVD that was observed (see supplementary data file 2). Code lists and operational definitions for each variable are provided in supplementary data file 1.

#### 2.3.5. Missing data

Missing data on systolic blood pressure, body mass index, non-HDL cholesterol, smoking status, ethnicity and index of multiple deprivation were imputed using multiple imputation by chained equations.^23,24^ All the other baseline predictors, exposure status at cohort entry date, an event indicator, and the Nelson-Aalen estimate of survival at the time of event/censoring, were used as predictors in the imputation model for each variable. Predictive mean matching was used as the imputation model for all missing variables. There were 10 imputation chains each of length 20. Performance of the imputation algorithm is evaluated and presented in supplementary material data file 2. This missing data approach corresponds with an implementation strategy of all data being available at implementation, which is our intention in the first instance.^25^

#### 2.3.6. Model development

Models were developed separately for male and female cohorts. The imputation procedure, and all steps in this section were done separately for both cohorts. After the imputation procedure, data was split into development (70%) and validation (30%) datasets. Imputing the entire dataset before splitting into development and validation dataset will result in an estimate of ideal model performance.^26^ Cross validation, or optimism adjusted bootstrap approaches, were not deemed necessary given the sample size.

The initial risk-estimation component fits a model which estimates the risk of CVD under the strategies ‘continue on current intervention strategy’. The model itself is a Cox proportional hazard model. Age, calendar time, systolic blood pressure, body mass index and non-HDL cholesterol were fitted as restricted cubic splines^27^ with 4 knots (age and calendar time) or 3 knots (all others) respectively. Knot locations were chosen manually for age at {25, 40, 57.5, 75}. Knot locations for the other predictors were data driven, by combining all imputed datasets, and calculating the 10^th^, 50^th^ and 90^th^ percentiles. All predictors except calendar time were interacted with the spline of age.

To estimate risk under ‘continue on current intervention strategy’, changes in treatments during follow-up that were observed in the development cohorts (“treatment drop-in”) needed to be addressed. We used an approach inspired by Xu et al.^17^ We created interval censored data at all time points where an individual started or stopped taking statins or antihypertensives, or changed their smoking status. Time varying versions of these variables were then created at each of these intervals, defined by their value relative to their value at baseline. The coefficients of these time varying variables were fixed to their causal average total effect, becoming offset terms. These effects were taken from literature^28–30^, converted to hazard ratios for model fitting,^31^ and are presented in Table 1. The effects of statins and antihypertensives were taken from an overview of systematic reviews of randomised controlled trials closely aligned with those used in the Millions Hearts Risk Assessment Tool.^32^ The effects of these interventions were assumed to be independent because no evidence was found to support any interactions. More details on the methodology for deriving the time-varying offset terms and estimating the values in Table 1 are provided in supplementary material data file 1.

**Table 1:** Treatment effects used in the baseline and intervention components, derived from the literature (see supplementary material data file 1), and expressed as odds ratios. Total effect is the effect of an intervention regardless of mechanism or pathway, which coincides with the usual ‘causal effect’ estimated in a trial. Direct effect is the effect of an intervention not via other measured factors. For example, the effect of smoking has indirect effects via BMI, SBP and cholesterol.

| <u>Variable</u> | <u>Causal odds ratio</u> | <u>Total or direct effect</u> |
| --- | --- | --- |
| Change in statin use during follow-up | 0.750 | Total |
| Change in antihypertensive use | 0.740 | Total |
| Initiates smoking during follow-up | 1.440 | Total |
| Stops smoking during follow-up | 0.800 | Total |
| Reduction in systolic blood pressure of 1mmHg | 0.974 | Direct |
| Reduction in Non-HDL cholesterol of 1 mmol/L | 0.812 | Direct |
| Reduction in BMI of 1 | 0.977 | Direct |
| Initiates smoking | 1.497 | Direct |
| Stops smoking | 0.851 | Direct |

#### 2.3.7. Sample size

A minimum required sample size was estimated using the criteria of Riley et al.,^33^ based on the mean follow up time and outcome incidence in the cohort, a conservative estimate of R2 = 0.15, 162 and 158 predictor parameters in the female and male models respectively, and a desired shrinkage of 0.99. The minimum required sample size was 437,350 in the female cohort, and 347,541 in the male cohort.

### 2.4. Development of intervention component with memory-based risk adjustment

The intervention component did not involve fitting any additional models. We assumed that interventions act through changes in modifiable risk factors (systolic blood pressure, body mass index, non-HDL cholesterol and smoking status). This approach can accommodate any intervention or combination of interventions, provided we can estimate its overall effect on each modifiable risk factor.

We created a tool which allows users to modify an individual’s current values for these risk factors. Based on these changes, we apply odds ratios to the risk (odds) estimated from the risk-estimation component. The causal odds ratios are shown in Table 1. Note that we use odds ratios here, whereas hazard ratios were required for the model fitting process in the initial risk estimation component.

We estimated the odds ratios using a causal directed acyclic graph (DAG^34^; Figure 2) and effect estimates from a variety of sources in the literature.^29,30,35–42^ This process is detailed by Jiang et al.^43^ Through this process we also estimated how starting statins or antihypertensives, or changing one modifiable risk factor, would affect the other modifiable risk factors (e.g. stopping smoking will on average result in an increase in body mass index and reduction in systolic blood pressure). This allows flexibility to estimate risk under a specific intervention, or under specified changes in the modifiable risk factors.

**FIGURE 2:**
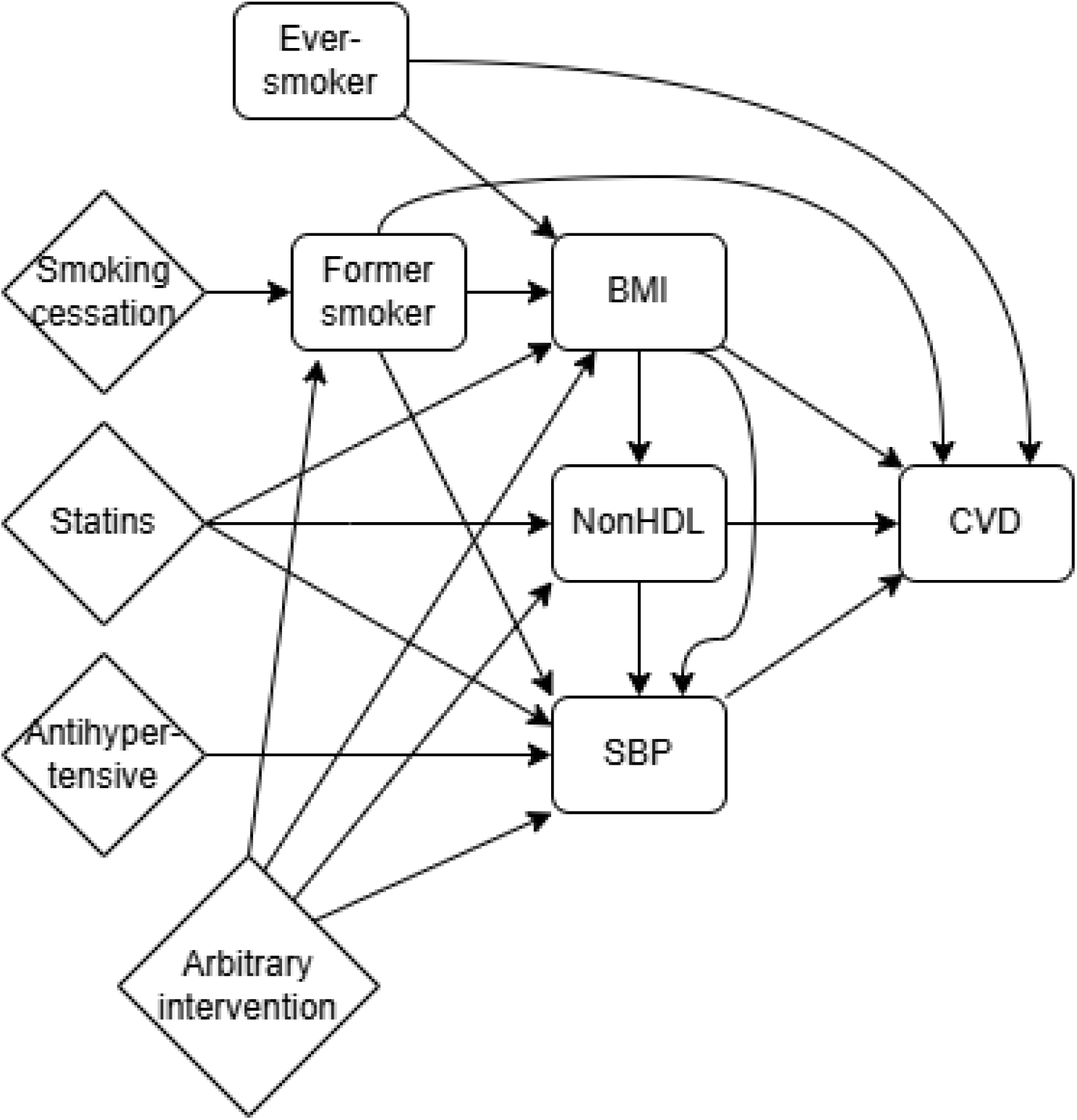
Directed Acyclic Graph of the causal structure. Modifiable risk factors are represented by rectangles. Interventions are represented by diamonds.

The model is designed to be used over multiple visits, following a similar process to that used in the Million Hearts Tool.^32^ At the first interaction (visit 0), the model estimates the individual’s risk using the initial risk estimation component and stores the values of the modifiable risk factors. Users can then explore hypothetical risk scores under different interventions, for example: What is my risk if I stop smoking? If I start statins? Or of I reduce my weight and blood pressure to healthy levels? These estimates can be based either on the total effect of an intervention, or on the effect estimates from the DAG combined with expected or targeted changes in each modifiable risk factor.

At each of follow-up visit, the following process is applied:

1. Remember the modifiable risk factor values from visit 0
2. Update other predictors (e.g. age, changes to medical history)
3. Estimate risk as if we had not intervened and modifiable risk factors remained at visit 0 levels,
4. Estimate current risk based on achieved levels of the modifiable risk factors
5. Estimate hypothetical future risks based on further interventions and modification of the risk factors.

An exemplar of how to use to model is given in section 3.1.

### 2.5. Model evaluation

Details of how evaluation was implemented across the 100 pairs of imputed development and validation datasets are provided in supplementary data file 1.

#### 2.5.1. Evaluation of the risk-estimation component

The chosen outputs for model evaluation were 10-year probabilities of CVD. We first evaluated the risk-estimation component. The 10-year risk of CVD for each individual in the validation cohort was estimated under intervention strategy of “continue on current interventions”. These are counterfactual risk predictions as many individuals do not follow this intervention strategy, i.e. they start or stop taking statins or antihypertensives or change smoking status during follow-up. There are a number of approaches to validating a prediction model which makes counterfactual predictions,^44–48^ however we again follow the work of Xu et al.^17^ This involves estimating counterfactual event times that would have been observed under the desired intervention strategy. The process for doing so is detailed in supplementary data file 1. Once the counterfactual survival times are estimated, model evaluation can proceed as normal.

Moderate calibration was assessed using calibration curves, the integrated calibration index, E50 and E90,^49^ and also the Kaplan-Meier observed risk vs mean predicted risk within 100 subgroups ranked by predicted risk.^50^ Discrimination was assessed using Harrell’s C-index.^27^ Decision curve analysis was not considered as further work is required to understand how to assess net benefit in the prediction under intervention context. Fairness was evaluated by estimating calibration and discrimination in subgroups defined by protected characteristics age, sex and ethnicity. We also evaluate performance by English region. Instability plots for individual risks were estimated.^51^

#### 2.5.2. Evaluation of the intervention component (temporal validation)

We evaluate the interventional part of the model by emulating the way in which the model is designed to be used in practice (section 2.5.3). We designate the index date defined in section 2.1 to be visit 0. We then extract cohorts at 1, 2, 3, 4 and 5 years after visit 0, referring to these as 1^st^, 2^nd^, 3^rd^, 4^th^ and 5^th^ follow-up visits, or ‘follow-up index dates’. All variables (predictors and exposures) are extracted in the same way as described in sections 2.3 and 2.4, but relative to the follow-up date, as opposed to the baseline index date. Individuals who have had a CVD event, or have been censored, prior to their follow-up index dates are excluded from the respective cohorts. When evaluating the interventional component, we choose both 5-year and 10-year risk because few individuals have sufficient follow-up at their fifth follow-up visit to allow us to evaluate 10-year risk predictions.

Next, for each modifiable risk factor we applied the following process, using systolic blood pressure as an example. We selected patients who had a non-missing value for systolic blood pressure at visit 0, and calculated the change in that modifiable risk factor between the first visit and follow-up visits. If there was no systolic blood pressure value recorded between visit 0 and the follow-up visit, we assumed that it was unchanged. We then estimated the risk of CVD using the risk-estimation component, where all variables (both exposures and predictors) except systolic blood pressure take the value extracted at the follow-up visit. For systolic blood pressure, we used the value from visit 0, and applied a relative risk based on its change between visit 0 and the follow-up visit. Calibration and discrimination were then estimated as described in section 2.6.1, including the estimation of counterfactual survival times. This process was also applied to all four modifiable risk factors simultaneously.

We also evaluated 5-year risk predictions estimated at these follow-up visits using only the risk-estimation component. This will help us understand whether any drop in performance discovered in the above analysis is driven by the interventional part of the model, or the fact there are temporal trends which we are not picking up. In these analyses, missing data in the follow-up visit cohorts was imputed using the value from the visit 0 cohort.

### 2.6. Patient and public involvement (PPI)

This study builds upon earlier PPI work, including a workshop with 19 members of the public, facilitated by 4 members of the Primary Care Research in Manchester Engagement Resource (PRIMER) and led by one of the current study’s co-authors (BM).^20^ Workshop members felt that a platform which encouraged a greater degree of interactivity with their NHS Health Check results using an online records access platform, could provide benefits to the NHS (by encouraging a healthy lifestyle, reducing NHS costs, saving GP time), and benefits to patients (e.g. through improved communication, by serving an educational role, providing additional motivation, and being less confrontational).^20^

### 2.7. Open Research

All analysis were conducted using R version 4.4.2. All code is available on the Manchester Predictive Healthcare Group GitHub page.^52^

## 3. Results

Table 2 contains the medical history and demographic information of our cohort at their baseline index dates. There was 301,383 (female) and 395,060 (male) outcome events respectively, and 210,543 (female) and 276,806 (male) events in the development cohorts. More information on total follow-up and event rates is available in supplementary data file 2.

**Table 2:** Baseline information.

| Characteristic | Male N = 9,510,838<sup>1</sup> | Female N = 9,899,565<sup>1</sup> |
| --- | --- | --- |
| Age |  |  |
| Median (5% Centile, Q1, Q3, 95% Centile) | 35 (18, 25, 48, 69) | 34 (18, 25, 49, 73) |
| N Missing (% Missing%) | 0 (0%) | 0 (0%) |
| Ethnicity |  |  |
| Bangladeshi | 68,082 (0.7%) | 61,655 (0.6%) |
| Black african | 229,698 (2.4%) | 260,426 (2.6%) |
| Black caribbean | 98,833 (1.0%) | 122,629 (1.2%) |
| Chinese | 109,615 (1.2%) | 164,065 (1.7%) |
| Indian | 267,095 (2.8%) | 262,903 (2.7%) |
| Other asian | 212,176 (2.2%) | 217,196 (2.2%) |
| Other ethnic group | 424,679 (4.5%) | 485,716 (4.9%) |
| Pakistani | 155,633 (1.6%) | 141,306 (1.4%) |
| White | 5,945,390 (63%) | 7,015,106 (71%) |
| Missing | 1,999,637 (21%) | 1,168,563 (12%) |
| Hypertension | 702,231 (7.4%) | 843,318 (8.5%) |
| Rheumatoid Arthritis | 22,677 (0.2%) | 59,199 (0.6%) |
| Atrial Fibrillation | 61,479 (0.6%) | 45,502 (0.5%) |
| Chronic Kidney Disease | 49,628 (0.5%) | 69,774 (0.7%) |
| Serious Mental Illness | 132,683 (1.4%) | 151,804 (1.5%) |
| Family History of CVD | 532,845 (5.6%) | 699,146 (7.1%) |
| Migraine | 271,571 (2.9%) | 701,889 (7.1%) |
| Systemic Lupus Erythematosus | 1,520 (<0.1%) | 11,851 (0.1%) |
| Diabetes |  |  |
| Absent | 9,225,275 (97%) | 9,663,232 (98%) |
| Type1 | 43,710 (0.5%) | 33,066 (0.3%) |
| Type2 | 241,853 (2.5%) | 203,267 (2.1%) |
| Impotence | 378,171 (4.0%) | 1,893 (<0.1%) |
| Corticosteroid Use | 43,066 (0.5%) | 71,647 (0.7%) |
| Atypical antipsychotic use* | 47,628 (0.5%) | 38,766 (0.4%) |
| Smoking status |  |  |
| Non-smoker | 4,013,333 (42%) | 5,287,522 (53%) |
| Ex-smoker | 1,740,158 (18%) | 1,927,811 (19%) |
| Current | 2,190,535 (23%) | 1,864,151 (19%) |
| Missing | 1,566,812 (16%) | 820,081 (8.3%) |
| Statin use at baseline | 337,046 (3.5%) | 319,790 (3.2%) |
| Antihypertensive use at baseline | 628,104 (6.6%) | 790,889 (8.0%) |
| Chronic obstructive pulmonary disease | 84,070 (0.9%) | 79,459 (0.8%) |
| Intellectual disability | 44,647 (0.5%) | 30,842 (0.3%) |
| Down syndrome | 5,187 (<0.1%) | 4,866 (<0.1%) |
| Oral cancer | 5,732 (<0.1%) | 3,320 (<0.1%) |
| Brain cancer | 3,106 (<0.1%) | 2,603 (<0.1%) |
| Lung cancer | 4,046 (<0.1%) | 3,744 (<0.1%) |
| Blood cancer | 22,943 (0.2%) | 19,544 (0.2%) |
| Pre-eclampsia | 20 (<0.1%) | 27,789 (0.3%) |
| Postnatal depression | 334 (<0.1%) | 130,832 (1.3%) |
| Body mass index |  |  |
| Median (5% Centile, Q1, Q3, 95% Centile) | 25.4 (19.8, 22.8, 28.5, 34.5) | 24.2 (19.1, 21.6, 28.3, 36.6) |
| N Missing (% Missing%) | 4,474,478 (47%) | 3,583,505 (36%) |
| Systolic blood pressure |  |  |
| Median (5% Centile, Q1, Q3, 95% Centile) | 130 (105, 120, 139, 155) | 120 (99, 110, 131, 150) |
| N Missing (% Missing%) | 3,972,552 (42%) | 2,042,261 (21%) |
| Non-HDL cholesterol |  |  |
| Median (5% Centile, Q1, Q3, 95% Centile) | 3.79 (2.11, 3.05, 4.50, 5.70) | 3.60 (2.10, 2.90, 4.40, 5.70) |
| N Missing (% Missing%) | 7,880,206 (83%) | 8,209,259 (83%) |
| Index of Multiple Deprivation (vintiles) |  |  |
| Median (5% Centile, Q1, Q3, 95% Centile) | 11.0 (2.0, 6.0, 15.0, 19.0) | 11.0 (2.0, 6.0, 15.0, 19.0) |
| N Missing (% Missing%) | 11,833 (0.1%) | 12,285 (0.1%) |

\*Second generation 'atypical' antipsychotic medication

Given the complexity of the fitted model, the coefficients cannot be presented in Tables. The model is provided as a freely available Rshiny application.^53^ The fitted model and cumulative baseline hazard are provided as R workspace objects (.rds) to support this Rshiny application, and can be downloaded from the CHARIOT GitHub repository.^52^

### 3.1. Evaluation of the risk estimation component

Figure 3 contains calibration plots of the models in the female and male cohorts. Both models were well calibrated across every combination of development and validation dataset. Calibration plots of the models within subgroups defined by geographical region, and protected characteristics ethnicity and age, as well as plots for the male cohort, and calibration metrics ICI, E50 and E90 are all provided in supplementary data file 2. The models were predominately well calibrated within subgroups defined by ethnicity, age and region. Both models have some calibration issues in the 17 – 30 age group. This is because a large number of individuals are under the age of 25. This is the position of our lowest knot, and below this point the effect of age is assumed to be linear (by the definition of restricted cubic splines^27^). However, given that individuals in this group are very low risk, this miscalibration is very small on the absolute scale. This is evident from the calibration metrics, where the E90 for this age group is smaller than many of the other age groups. There is also some under-prediction of risk among higher risk individuals in the North West.

**Figure 3:**
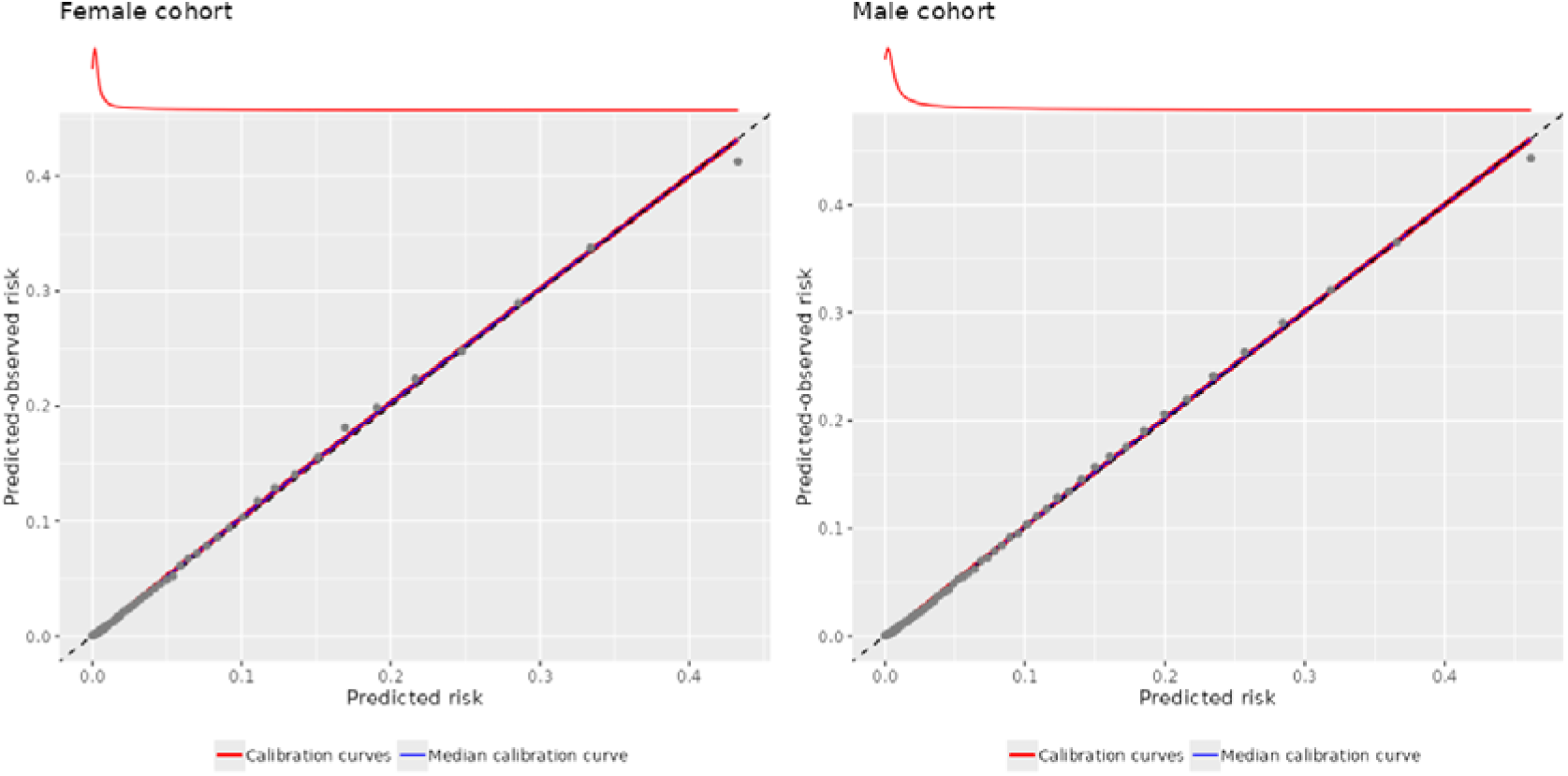
Calibration plots. Smoothed calibration curve, and binned calibration plot with centiles or predicted risk.

The discrimination in the entire validation cohort (Harrel’s C-index), and subgroups defined by protected characteristics are presented in Figure 4. Confidence intervals were estimated but are too small to be visible in the plots. The C-index is 0.87 for women and 0.86 for men. Discrimination remains similar within subgroups defined by ethnicity or region. As expected, there is a considerable drop in performance within age groups, because age is a strong prognostic factor that interacts with all other predictors. Instability plots of the model,^51^ plotted for 3,000 randomly selected individuals, are presented in supplementary data file 2.

**Figure 4:**
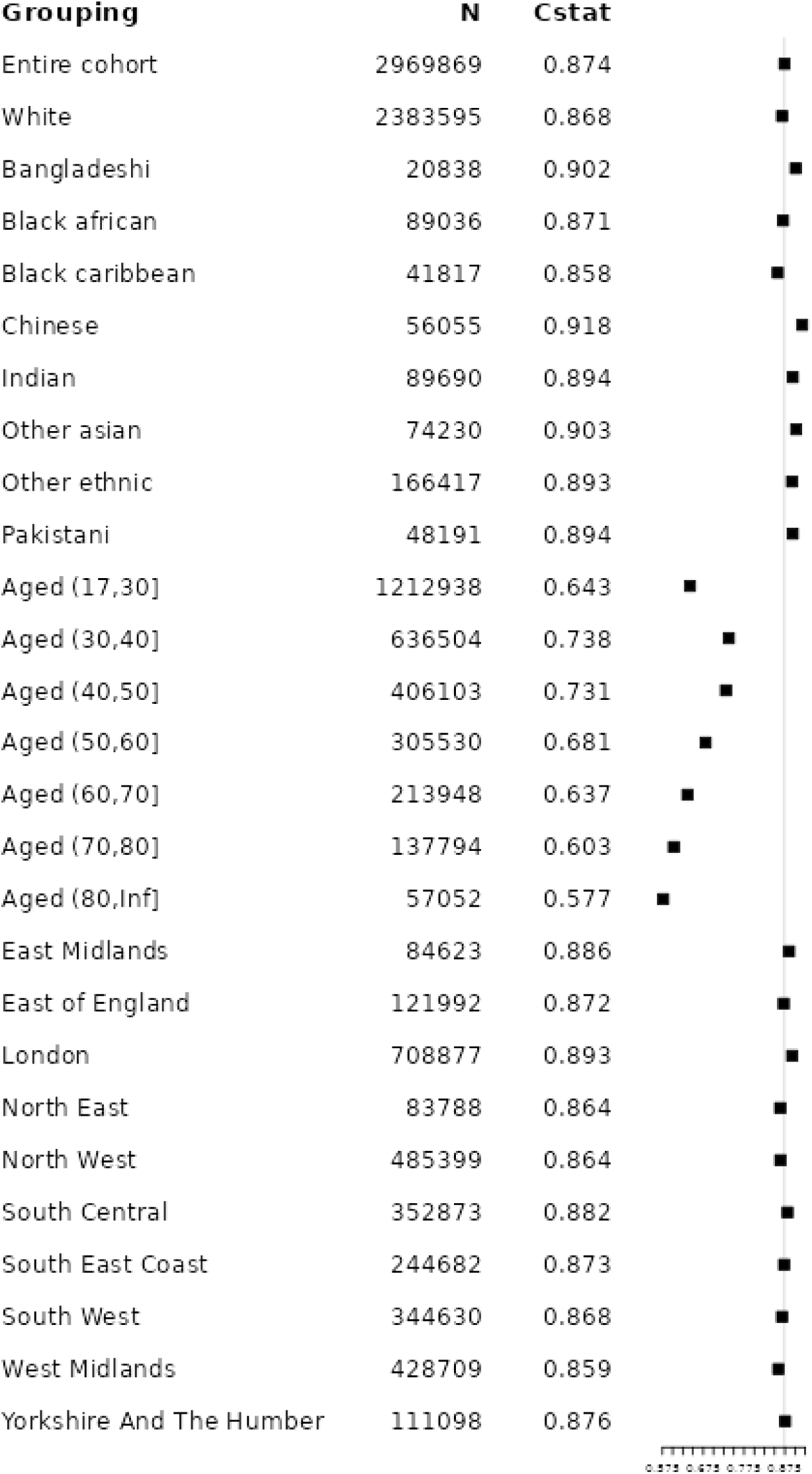
Discrimination of the model in the entire cohort and within subgroups defined by protected characteristics.

### 3.2. Evaluation of the intervention component (temporal validation)

Figure 5 contains the calibration plots of the initial risk estimation component predicting 5-year risk at 1, 3 and 5 year follow-up visits among females. It is evident that the further away from visit 0 the index date is defined, the model begins to over-predict in the high-risk individuals. This could be due to selection bias, as unhealthy individuals will leave the cohort (either through having CVD events or dying), and no new individuals are added to the cohort. Plots for the male cohort and evaluating 10-year predictions, and discrimination, are provided in supplementary data file 2. Conclusions are broadly similar.

**Figure 5:**
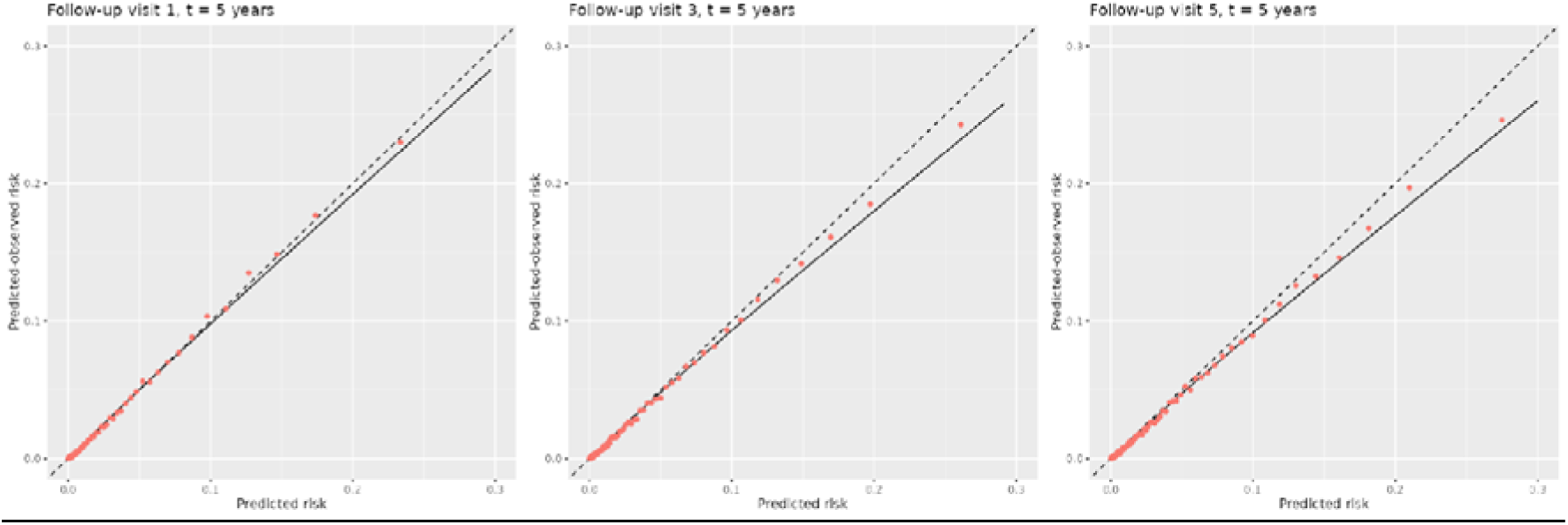
Calibration plots of the initial risk estimation component predicting 5-year risk at 1, 3 and 5-year follow-up visits, female cohort.

Figure 6 contains the calibration plots of the intervention component predicting 5-year risk at the 1-year follow-up visit, for modification of each of the modifiable risk factors. The calibration of the intervention component when modifying non-HDL cholesterol, body mass index and smoking status remains quite strong. There is overprediction of higher risk individuals for systolic blood pressure. Plots for the male cohort, evaluating 10-year predictions, and making predictions at the 2, 3, 4 and 5-year follow-up visits, and discrimination, are provided in supplementary data file 2. Conclusions are broadly similar.

**Figure 6:**
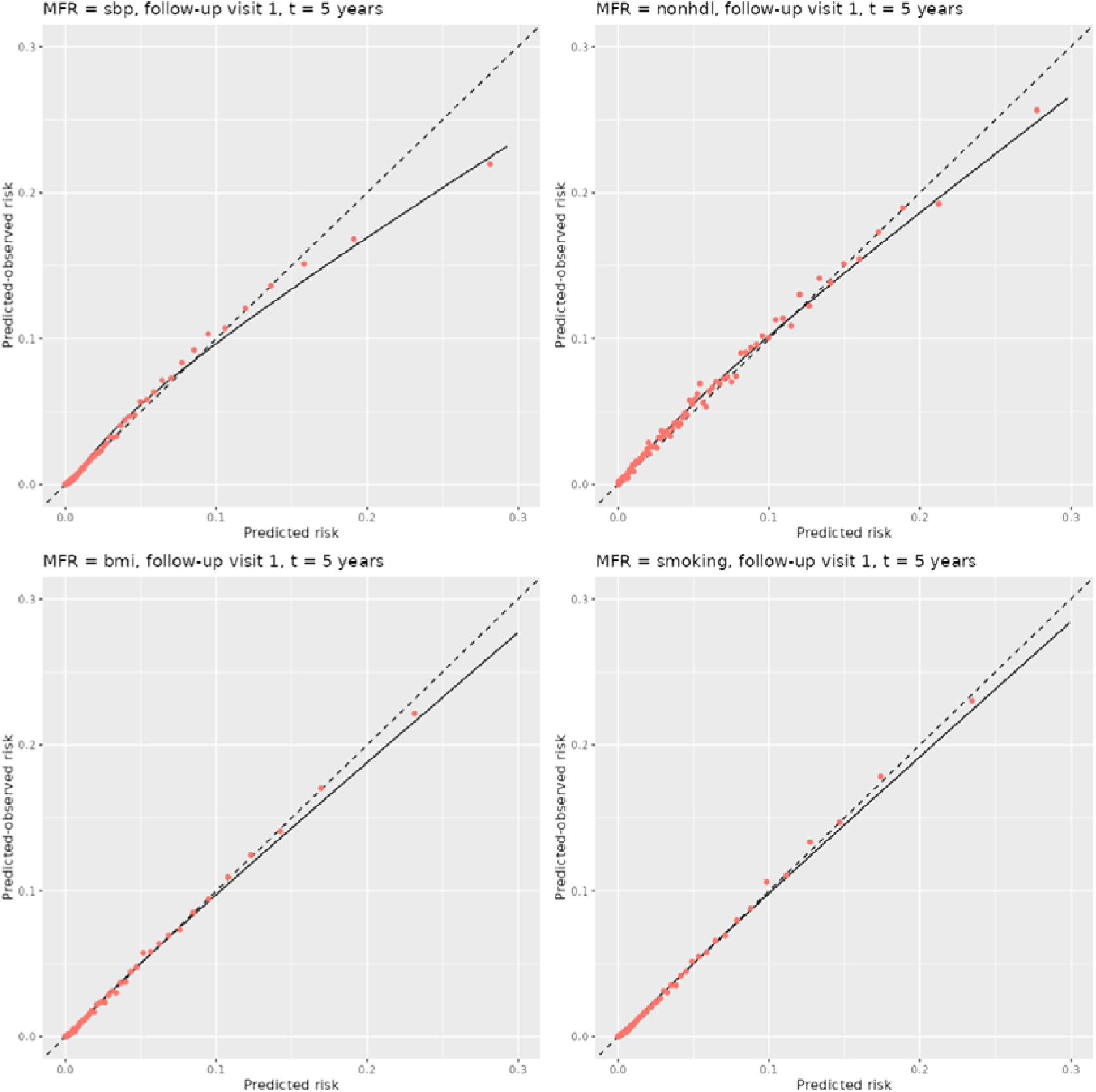
Calibration plots of the intervention component for each of the modifiable risk factors at the 1-year follow-up visit, female cohort.

### 3.3. Worked example

#### Setting

To illustrate CHARIOT’s clinical utility, we create a scenario similar to that used to showcase the Million Hearts Tool. We consider a 70-year-old Black Caribbean woman, a current smoker with elevated risk factors: systolic blood pressure 160mmHg, BMI 30 (165cm, 81.68kg), and non-HDL cholesterol 7 mmol/L. She has no other comorbidities and is not on statins or antihypertensives but has a poor diet and does not exercise regularly.

#### Visit 0

The individual attends an NHS health check. Using CHARIOT, her 10-year CVD risk without intervention is 18.97%.

The clinician discusses risk reduction strategies: statins, antihypertensives, smoking cessation, or dietary/exercise changes. To inform this discussion, we estimate 10-year risk under various interventions (Table 3).

**Table 3:** Predicted 10-year CVD risk under a range of interventions, and hypothetical reductions in modifiable risk factors.

| **Intervention** | **Predicted 10-year CVD risk under intervention** |
| --- | --- |
| Statin therapy | 14.32% |
| Antihypertensive therapy, reducing SBP by 5 mmHg | 17.03% |
| Antihypertensive therapy, reducing SBP by 10 mmHg | 15.26% |
| Antihypertensive therapy, reducing SBP by 15 mmHg | 13.64% |
| Smoking cessation | 15.77% |
| Weight loss of 5kg | 17.24% |
| Weight loss of 10kg | 15.64% |
| Combination therapy (reduces Non-HDL by 2mmol/L, SBP by 20mmHg, and weight by 10kg) | 5.49% |

The individual prefers avoiding medication unless necessary and is interested in a smoking cessation programme, which could have broader health benefits. She’s willing to attempt dietary and exercise changes but is uncertain about sustaining them. The predicted risks show smoking cessation offers similar benefit to statins or antihypertensives. Given her motivation for smoking cessation, she enrols in a programme and agrees to attempt to dietary improvements and increased physical activity (via the NHS weight loss plan), aiming to reduce risk below 15.77%. A follow-up is scheduled for one year to reassess medication needs.

#### Follow-up visit 1

One year later, the individual has successfully stopped smoking and improved her diet, reducing non-HDL cholesterol by 2 mmol/L and losing 5kg. However, she struggled to increase physical activity, and blood pressure remains unchanged. Following the process in section 2.5.3:

First, we re-estimate baseline risk one year on, updating non-modifiable factors (e.g., age increased to 71; potential new comorbidities like COPD or new medications like corticosteroids). Modifiable risk factors remain at visit 0 values, estimating 10-year CVD risk without intervention: 20.05%.

Next, we estimate current risk based on achieved modifiable risk factor levels. With reduced BMI, improved cholesterol, and smoking cessation, her current 10-year risk is 11.89%.

A new discussion begins regarding further treatment. Given successful cholesterol reduction and willingness to continue dietary changes, statins are deemed unnecessary. With persistent high blood pressure, we estimate risks under antihypertensive therapy achieving reductions of 5mmHg (10.58%), 10mmHg (9.41%), 15mmHg (8.35%), or 20mmHg (7.4%). The individual agrees to antihypertensives. At follow-up visit 2, the same process from section 2.5.3 applies.

## 4. Discussion

### 4.1. Interpretation

We have developed a clinical prediction model for CVD that moves beyond risk identification toward genuine clinical decision support. Rather than simply flagging that a patient is high risk, CHARIOT enables a personalised conversation: what would happen to this patient’s risk if they started a statin, stopped smoking, or made sustained lifestyle changes? This is an advance over both classical and transformer based CVD prediction models which focus on observed risks and are agnostic to intervention.^2–8,54^ Current risk assessment approaches in England –centred on QRISK and the NHS Health Check– are designed to support the initiation of statin therapy rather than ongoing treatment monitoring.^2,11^ CHARIOT supports sustained engagement with prevention across multiple clinical encounters, enabling re-evaluation of treatment options as a patient’s risk factors change over time. This is a key step towards personalised preventative care that digital health infrastructure increasingly makes possible, but that risk tools have not yet delivered.

Alongside the novel prediction under intervention aspects, the model preserves good predictive performance with respect to traditional metrics - it is well calibrated and has discrimination meeting or exceeding the discrimination of models currently used to guide clinical care around CVD.

With regards to fairness, the models are well calibrated and discriminate well in subgroups defined by ethnicity and region in England. There is a considerable drop in discrimination within subgroups defined by age compared with performance in the entire validation cohort; this highlights the importance of age for predicting CVD risk.

### 4.2. Comparison with existing literature

We are aware of three other models developed for prediction of CVD under intervention. The Million Hearts Risk Assessment Tool^32^ considers statin therapy, blood pressure lowering medication, aspirin, smoking cessation and combinations of these interventions. The PEER Simplified Cardiovascular Decision Aid Tool^55^ considers statins, blood pressure lowering, Ezetimibe, proprotein convertase subtilisin/kexin type inhibitors and Fibrates medications, mediterranean diet and physical activity. Qintervention^56^ considers statins, reducing body mass index to 25, reducing systolic blood pressure to 140mmHg, and stopping smoking. CHARIOT is an advance over all of these models because it develops the baseline risk estimation model using causal inference techniques to address treatment drop-in, and decomposes intervention effects into direct and indirect components, allowing for prediction under arbitrary combinations of interventions rather than just pre-defined options. Equally importantly, none of these tools have been designed with repeated digital interactions in mind, either for use across multiple clinical visits or through patient-facing health record platforms. CHARIOT’ s memory property and multi-visit architecture address this gap.

Our approach is most similar to the Millions Hearts Tool, which conducted a systematic review of the literature to obtain effect estimates of risk reducing therapies and applied these to baseline risks derived from the 2013 Pooled Cohort Equations.^5,6^ The Million Hearts Tool has the same memory property and is implemented in the same way at follow-up visits. CHARIOT has developed a new model for the initial risk estimation component, as opposed to using an existing model. This has enabled the model to be developed in a large cohort representative of the target population (with noted limitations for implementation in Wales, Scotland and Northern Ireland due to available linked data), with strong discriminative performance. CHARIOT has also used causal inference techniques to deal with ‘treatment drop-in’^16,17,19,57^) during the development of the initial risk estimation component, although the impact of this in a situation where individuals are both on and off treatment at baseline needs to be explored in more detail. The effect estimates for the interventions and changes in modifiable risk factors are similar, with one notable exception. CHARIOT has assumed a relative risk reduction of 0.8 per 10mmHg reduction in systolic blood pressure (taken from a 2016 meta-analysis^58^), compared to the 0.65 (taken from a 2009 systematic review^59^) used in the Million Hearts Tool.^32^ This is by no means an exact science, and inconsistencies were found when trying to validate this choice.^43^ Another important difference is that CHARIOT decomposed the effect of interventions into direct and indirect effects, allowing the prediction of risk under arbitrary interventions, with hypothesised effects on each of the modifiable risk factors. While this flexibility is an advantage, this process required utilising a range of data sources including causal mediation and mendelian randomisation studies, which should be considered weaker evidence. Finally, to avoid over-estimating treatment effects, the Million Hearts Tool implemented floor values, below which the predictions under intervention couldn’t go, which were based on the predicted risk of untreated optimal levels of all risk factors at the same age, sex and race. This approach will be considered in future iterations of CHARIOT.

The PEER Simplified Cardiovascular Decision Aid^55,60^ estimates baseline risk from a range of different models depending on your location, and Qintervention from QRISK2.^61^ Both combine these with effect estimates of risk reducing therapies, however do not consider combinations of interventions, are restricted to a pre-defined list of interventions, do not account for changes in treatment or treatment drop-in^16,17,19,57^ during model development, or have a well-defined way to be used at follow-up visits.

### 4.3. Strengths, limitations and assumptions

The CHARIOT model has been developed in a large cohort representative of the target population and has strong performance. It has been designed with a specific clinical interaction in mind,^62^ and has a memory property to allow it to be used at follow-up visits for ongoing treatment. The approach combines the strengths of different data sources, routinely collected electronic health record data for estimating the baseline risk, and effect estimates from a mix of trials and mendelian randomisation studies for intervention component.

The combination of different data sources could, however, be a weakness as the effect estimates come from studies with different cohort definitions, outcome definitions and modelling approaches. While it would be preferable to estimate these in a consistent manner, this is not possible given the desire to use the methodological strength of trials carried out on statin and antihypertensive treatment. Instead, we must aim to combine this information in the best way possible.^63^ This also has implications for estimating uncertainty intervals,^35^ as it is still an open question how we propagate the uncertainty from the estimation of the causal effects into the risk scores. We have estimated stability plots for the risk scores from the initial risk-estimation component (supplementary data file 2).

A second limitation is that lifestyle interventions may reduce CVD beyond reductions in the modifiable risk factors we have considered, for example through heart muscle, mental state or hormones. One study showed that exercise could operate through improvements to insulin sensitivity, improved lining of blood vessels, reduced heart rate and increased antioxidants, none of which are routinely collected or recorded in the primary care electronic health record.^65^ We may therefore be underestimating the benefit for some interventions. A third limitation is that we assume that reducing one of the modifiable risk factors (i.e. systolic blood pressure from 140 to 120) has the same effect on CVD, irrespective of how that reduction was achieved (consistency assumption). The impact of this assumption or whether it holds is unclear. To mitigate the assumption of positivity, we do not allow the model to predict risk under interventions which an individual would not be eligible for (e.g. statins for individuals with risks under 7.5%). A fourth limitation is that while the ideal target population is the entire UK, due to data restrictions, the model could only be developed in English individuals. Validation and possibly model updating would therefore be necessary in the other UK nations. Also, while using CPRD has many advantages, the nature of electronic health records means there is likely measurement error and the analysis is ultimately dependent on the choice of code lists. Further validation may help assess the impact of this.

### 4.4. Clinical implications and implementation

CHARIOT is designed for implementation in two complementary contexts: discussions between healthcare professionals and patients, for example, during the NHS health check, and use by patients using self-care apps that access their electronic healthcare records.

These contexts are not mutually exclusive: the same underlying model can support both a GP discussing statin initiation with a patient, and that patient later exploring their own risk and potential interventions through an online portal.

The alignment with response efficacy theory is important here. Evidence suggests that patients are more likely to initiate and sustain behaviour change when they understand their personal risk and the benefit of acting.^18,66^ Generic risk scores do not provide this, while CHARIOT does. Whether this translates into measurable improvements in behaviour change and clinical outcomes in practice remains to be tested, and is a priority for future research.

From an implementation perspective, the model’s memory property (its ability to recall baseline risk factor values and update predictions at follow-up visits) is both a clinical strength and a practical consideration. Realising this functionality in electronic health record systems will require those baseline values to be captured and stored in a structured, retrievable way.

The back-end of this algorithm is publicly available as an Rshiny application,^53^ demonstrating feasibility. A front-end application is in development.^67^ Translation into production clinical systems will require co-design with clinicians, patients, and health informatics teams to ensure the tool is usable and trusted in real-world settings.

### 4.5. Future work

A key area for future work is to extend the methodology to the competing risk setting. This would allow us to predict lifetime risk, and the risk of multiple outcomes. Being able to estimate the risk reduction of a lifestyle change on a number of potential outcomes (e.g. CVD, type 2 diabetes and chronic kidney disease) may be more powerful in motivating behaviour change. It also allows the end user to make decisions based on a more holistic view of their health. Similarly, being able to estimate the reduction in lifetime risk under an intervention may be able to help motivate behaviour change in younger individuals, whose 10-year risk will be very small. Understanding how to predict under intervention in a competing risk setting is essential to achieve both estimation of lifetime risk and multiple outcomes. We will explore the feasibility of incorporating CHARIOT into clinical EHR systems and patient-facing records access platforms to facilitate discussions around CVD risk reduction, and help motivate health behaviour change.

### 4.6. Conclusion

CHARIOT predicts future CVD risk under interventions which modify systolic blood pressure, body mass index, non-HDL cholesterol and smoking status, whilst retaining strong predictive performance compared to existing models.

## Data Availability

Operational definitions for all variables and the process for extraction of the CPRD data are provided in supplementary data file 1. All code and codelists are available on the CHARIOT GitHub repository. The code supporting the extraction of the CPRD data has been written into a freely available R package, rcprd.66 CPRD data are not publicly available. Details of the application process and conditions of access are available at: https://www.cprd.com/data-access. The model developed in this paper has been made publicly available as an Rshiny app: https://alexpate30.shinyapps.io/rshiny_chariot_prototype3/

https://github.com/manchester-predictive-healthcare-group/CHI-CHARIOT/

## 6. Supporting statements

### 6.1. Supplementary data files

Supplementary data file 1: methods and technical appendix Supplementary data file 2: supplementary tables and figures Supplementary data file 3: TRIPOD+AI checklist

### 6.2. Availability of code and materials

Operational definitions for all variables and the process for extraction of the CPRD data are provided in supplementary data file 1. All code and codelists are available on the CHARIOT GitHub repository: https://github.com/manchester-predictive-healthcare-group/CHI-CHARIOT/tree/main/generic-data-extraction. The code supporting the extraction of the CPRD data has been written into a freely available R package, rcprd: https://CRAN.R-project.org/package=rcprd. CPRD data are not publicly available. Details of the application process and conditions of access are available at: https://www.cprd.com/data-access. The model developed in this paper has been made publicly available as an Rshiny app: https://alexpate30.shinyapps.io/rshiny_chariot_prototype3/. A study protocol was not prepared and this study was not registered.

### 6.4. Ethics and consent to participate

CPRD has ethics approval from the Health Research Authority to support research using anonymised patient data. CHARIOTS application (protocol 22_002333) was reviewed via the CPRD Research Data Governance (RDG) process to ensure that the proposed research is of benefit to patients and public health. The data that CPRD receives from GP practices is pseudonymised at source, meaning that GPs do not need to seek individual patient consent when they share data with CPRD, however patients are able to opt out.

### 6.6. Author contributions

BM, MS and NP conceived the work. AP, BJ, YH, BM and MS designed the work. AP, BJ, YH, BM and MS acquired data. AP and BJ implemented analyses and interpreted results with YH, SG, DS, NP, BM and MS. AP drafted the manuscript which was then substantively revised by BJ, YH, SG, DS, NP, BM and MS.

## 6.7. Acknowledgments

Thank you to the University of Manchester Research IT team who maintain the computational shared facility on which these analyses were run.

An LLM was used for making text more concise in section 3.3 The authors have reviewed and take final responsibility for the final text.

