## Supplementary data files 1, 2 and 3 for "Predicting cardiovascular risk under intervention: Development and internal validation of the CHARIOT Model in 19 million adults": CHARIOT applied - Supplementary data file 1 - methods.docx

### 1. Code for CPRD Aurum data extraction

The code and supporting documentation for the CPRD Aurum data extraction is available on GitHub (see folder generic-data-etraction). This includes operational definitions for each variable extracted, the code lists used to define each variable, name and description of all code used for data extraction, and information on the structure of the code. The code used for data extraction in this study formed the basis for development of an R package, rCPRD,^1^ ‘to simplify the extraction and processing of CPRD Aurum data, and creating analysis-ready datasets’. If you are reading this data extraction document to better understand how to extract and work with CPRD Aurum data, you are better off looking at the package documentation. Specifically, the rCPRD package vignette: <https://alexpate30.github.io/rcprd/articles/rcprd.html>, is a step-by-step guide on how to use this package to work with CPRD AURUM data, and will be kept up to date. The code and associated information used for the data extraction in this project is available in the generic data extraction GitHub repo.^2^

### 2. Code for running analyses

This section documents the code for running the analyses in the associated manuscript. All analyses were run in R version 4.4.2.

In order to re-use the code, the working directory should be set to the folder containing the ‘code’ subfolder. This must be set using setwd() at the top of every .R file. Sadly, we could not find a way to get the ‘here’ package working with the computational facility and data storage requirements we had.

In order to re-use the code, there should also be a ‘data’, ‘R, and ‘figures’ folders in the same directory.

The ‘data’ and ‘figures’ folders should contain a subfolder ‘p4’, in which many data objects will be stored. The ‘R’ folder contains functions that have been written to be used repeatedly (e.g. estimating calibration plots, augmenting data, etc).

The code itself is then all contained within a subfolder ‘p4’. Folders p1-p3 contain code from previous versions of the algorithm which have been omitted.

Inside ‘code’, there are then 8 subfolders, which we detail the contents of here:

**p1_data_prep:** data preparation and imputation.

**p2_data_prep_further_mfr:** further data preparation, to create interval censored data with respect to changes in smoking status, antihypertensive use and statin use.

**p3_devel:** Development of the clinical prediction model.

**p4_validate:** Internal validation of the clinical prediction model (calibration, discrimination, and instability plots). This is the validation of the ‘risk estimation layer’.

**p5_validate_temporal:** Temporal validation. This is the validation of the ‘intervention layer’.

**p6_validate_supplementary:** Validation of a version of the clinical prediction model where we fixed the effect of the modifiable risk factors, rather than splitting out the ‘risk estimation’ and ‘intervention’ layers.

**p7_shiny:** code for the rshiny app.

**p8_collate_results:** Collate all tables, figures, and generate documents to be used for supplementary material.

Within each of these folders, .R files are all prefixed with ‘p1’, ‘p2’, …., and are designed to be run in order. Often (but not always), subsequent .R files are dependent on objects created in previous .R files.

#### 2.1. Dictionary of all files

If a file is not mentioned here, it is surplus to requirements or used for exploration of the data.

##### 2.1.1. p1_data_prep

p1_extract_variable_components.R: Extract components of composite variables.

P2_create_cohort.R: Create baseline cohort.

P2_table1.Rmd: Create table of baseline cohort.

P3_create_variable_objects.R:

P4_impute_cohort_prototype3.R: Multiple imputation

P5_create_imp_comb.R: Combine the parallelised imputation objects into a single ‘mids’ object.

P6_assess_imputations.R: create density and convergence plots

P7_create_split_sample.R: create development and validation datasets

P8_get_knot_locations.R: estimate knot locations

P9_calculate_HR_OR_minimax: Convert effect estimates into hazard ratios for applying the offset approach.

##### 2.1.2. p1_data_prep_further_mfr

p1.1_extract_modifiable_risk_factors.R: query SQLite database for test records on BMI, SBP, Non-HDL cholesterol and smoking status.

p1.2_extract_prescriptions.R: query SQLite database for statin and antihypertensive prescriptions.

p3.1A_extract_smoking_followup_for_model.R: Create interval censored data based on when an individual changes smoking status. NB: this variable denotes an individuals smoking status, NOT, their smoking status relative to smoking status at baseline, which is what is required to adjust for changes in status during followup. This is because smoking status has missing data at baseline, and therefore this step is applied later.

p3.1B_combine_smoking_followup_for_model.R: Combine parallelised files from 3.1A.

p3.2A_extract_prescriptions_followup_for_model.R: Create interval censored data based on when an individual changes statin or antihypertensive status. (0 = off treatment, 1 = on treatment).

p3.2B_combine_ prescriptions_followup_for_model.R: Combine parallelised files from 3.2A.

p3.2C_augment_ prescriptions_followup_for_model.R: Augment file from 3.2B, to be a -1, 0, 1 variable, relative to the individuals status at baseline.

p4.1_layer_cohort_split_times.R: layer the interval censored data from each of smoking status, statins and antihypertensives, to create one dataset with intervals when any of these time-varying variables changes.

P4.2_combine_layer_cohort_split_times.R: the code from p4.1 is designed to be parallelised. This program combines into one file.

##### 2.1.3. p3_devel

p1_prototype3.R: Fit the model with 3 knots for age (40, 57.5, 75)

p1_prototype3_4knots.R: Fit the model with 4 knots for age (25, 40, 57.5, 75)

p1_prototype3_4knots_caltime.R: Fit the model with 4 knots for age (25, 40, 57.5, 75), where calendar time is also adjusted for as a predictor.

p3_prototype3_assess_nonlinear.R: Create plots for the shape of the interaction between age and all other predictors. Same for each of the models fitted in p1.

p5_worked_exemplar.Rmd: worked exemplar for the manuscript.

p6_sampsize_calc.R: sample size calculation.

P7_tests_for_functions_to_estimate_risks.R: tests to check some user written functions are working properly.

##### 2.1.4. p4_validate

p1_prototype3_calculate_cf_surv_times.R: Calculate counter factual survival times for the model with 3 knots for age.

p1_prototype3_calculate_cf_surv_times_4knots.R: Calculate counter factual survival times for the model with 4 knots for age.

P2_prototype3_calibration_ph_d1v1.R: Estimate calibration plot for 1 model in 1 validation dataset (there are 10 of each given the multiple imputation).

P2_prototype3_calibration_ph_MxM.R: Estimate calibration plot for every pair of model and validation dataset (there are 100 given the multiple imputation), estimate median, 2.5^th^ and p7.5^th^ percentiles.

P3_prototype3_discrimination_d1v1.R: Estimate discrimination for 1 model in 1 validation dataset (there are 10 of each given the multiple imputation).

P3_prototype3_discrimination_MxM.R: Estimate discrimination for every pair of model and validation dataset (there are 100 given the multiple imputation), estimate median, 2.5^th^ and p7.5^th^ percentiles.

P4.1_instability_fit_models.R: Fit models with 3 knots for age in bootstrapped datasets in order to estimate instability plots.

P4.1_instability_fit_models_4knots.R: Fit models with 4 knots for age in bootstrapped datasets in order to estimate instability plots.

P4.1_instability_fit_models_4knots.R: Fit models with 4 knots for age and calendar time included as a predictor in bootstrapped datasets in order to estimate instability plots.

P4.2_instability_plots_individual_risks.R: Create instability plots

##### 2.1.5. p5_validate_temporal

p1.1_temporalv_combine_cohorts.R: Combine cohorts at 1/2/3/4/5 years post follow-up into a single dataset.

p1.2_temporalv_create_individual_cohorts: Create individuals cohorts at 1/2/3/4/5 years post follow-up, fill in missing data, and give a common naming convention.

p2_temporalv_layer_cohort_split_times.R: Get the interval censored data relative to the new values of treatment status (statins, antihypertensives and smoking status) at the follow-up index dates.

p4_temporalv_prototype3_calculate_cf_surv_times.R: estimate counterfactual survival times (similarly named programs for models developed with 4 knots for age and calendar time included as a predictor).

p5_temporalv_initial_layer_prototype3_d1v1.R: temporal calibration of the initial risk estimation layer.

p6_temporalv_intervention_layer_prototype3_d1v1.R: temporal calibration of the intervention layer.

##### 2.1.6. p7_rshiny

create_shiny_data.R: create of objects for the rshiny app.

app.R: the rshiny app

##### 2.1.7. p8_collate_results

p1_collate_tables_word.Rmd: Collate all tables into a word document.

p2_collate_figures_and_tables.Rmd: Collate all figures and tables into a html file.

p3_compare_models_age_knots.Rmd: Compare models with 3 and 4 knots for age.

P4_compare_models_caltime.Rmd: Compare models with 4 knots for age, but calendar time included or not included as a predictor.

#### 2.2. Required changes to implement code

**TL;DR**: Finally, the code has been written on the assumption that the CPRD Aurum data has already been extracted, formatted into required R .rds objects. The code is therefore not fully re-usable without some minor edits required, which are detailed here. These edits are all required in the data preparation folders (p1_data_prep) and (p2_data_prep_further_mfr).

##### 2.2.1. Reading in the baseline cohort file

Extraction of the CPRD data used in this project was undertaken to support a number of different analyses. Therefore, the data and analysis cohort were extracted and stored in a distinct place from the directory system (‘code’, ‘data’, ‘figures’, ‘R’) described above.

The cohort itself is a data.frame, stored as an .rds file, with one row per individual, and one column for each variable. This contains information about each individual extracted at their baseline index date (latest of: the start of the study period (1/1/2005), attaining at least one year of registration with a contributing practice to CPRD Aurum, attaining age 18). The structure of the data.frame, including variable names and formats, is provided in aaa_cohort_file_structre.txt. The exact process for the data extraction used in this study, and associated code is detailed in the data extraction GitHub page.^2^ The code written to undertake this cohort extraction has been expanded into an R package, rcprd.^17^

This cohort must be read in and saved into the workspace within which all this code operates. Specifically, in the file: ‘*p1_data_prep/p2_create_cohort.R’*, at line 21, the ‘*readRDS()*’ command should be changed to read in the cohort file from wherever it is stored

Also note, the file: ‘p1_data_prep/p1_extract_variable_components.R’ extracts the components of composite variables (e.g. BMI, Non-HDL cholesterol, etc). This was done for exploratory reasons and is not required for the fitting of this model. It also requires querying an SQLite database (see rcprd^17^ and the data extraction GitHub page^2^), so we do not recommend running this file. It has been provided for transparency, as the file ‘*p1_data_prep/p2_create_cohort.R’* depends on it. In the file ‘*p1_data_prep/p2_create_cohort.R’* the code which reads in the variable components and merges it with the cohort (i.e. line 34 – line 80) can be ignored. The goal of this program is to read in the pre-existing cohort file and save into the working direction structure for this project.

**In summary**

We recommend not running ‘*p1_data_prep/p1_extract_variable_components.R*’, and then simply read in the cohort file which should already have been created, re-format the smoking variable, and save the cohort to “data/p4/cohort_prototype3.rds”.

##### 2.2.2. Reading in the cohort files at follow-up index dates

Similarly to section 3.1, the cohorts at the follow-up index dates are created outside of the directory system provided here. The following programs read these cohorts in and must be amended accordingly:

‘*p5_validate_temporal/p1.1_temporalv_combine_cohorts.R*’ (line 26) ‘*p5_validate_temporal/p1.2_temporalv_create_individual_cohorts.R*’ (line 70)

The exact process for the data extraction used in this study, and associated code, is detailed in the data extraction GitHub page.^2^ The code written to undertake this cohort extraction has been expanded into an R package.^17^

##### 2.2.3. Querying an SQLite database to extract prescription and smoking information during follow-up

To fit the models in the associated manuscript and estimate counterfactual survival times, we needed to extract internal censored data, based on changes in smoking status, statin use and antihypertensive use during an individual’s follow-up. We also wanted to extract systolic blood pressure, body mass index and Non-HDL cholesterol during follow-up, to validate the interventional part of the model. This required some extra data extraction on top of the cohort described in section 3.1 of this guide.

The CPRD data was extracted and stored in an SQLite database for efficient querying. As described in section 3.1, this was stored in a different directory structure as it was being used for multiple analyses. This SQLite database is queried in programs:

*‘p2_data_prep_further_mfr/p1.1_extract_modifiable_risk_factors.R’*

*‘p2_data_prep_further_mfr/p1.2_extract_prescriptions.R’*

This requires the reading in of codelists, and querying of the SQLite database, both are which are stored outside of the previously specified directory structure (‘code’, ‘data’, ‘figures’, ‘R’).

The database queries are then saved as .rds files for easier access. Based off these queries, the interval censored data is then created through data manipulation in programs:

*‘p2_data_prep_further_mfr/3.1A_extract_smoking_followup_for_model.R’*

*‘p2_data_prep_further_mfr/3.1B_combine_smoking_followup_for_model.R’*

*‘p2_data_prep_further_mfr/3.2A_extract_prescriptions_followup_for_model.R’*

*‘p2_data_prep_further_mfr/3.2B_combine_ prescriptions_followup_for_model.R’*

*‘p2_data_prep_further_mfr/3.2C_augment_ prescriptions_followup_for_model.R’*

##### 2.2.4. Parallelisation and command line inputs

Due to the large sample sizes used in this study (~ 20 million in total), there are a number of files which were written so they could be run on a computational cluster and parallelised. In order to parallelise, the task ID is read in at the command line.

Some files a gender (1/2) variable is read in at the command line.

Some files a medication (‘statin’ or ‘antihypertensive’) is read in at the command line.

These files are listed here:

p1_data_prep/p4_impute_cohort_prototype3.R (parallelisation over task ID 1 - 10, and male/female).

p2_data_prep_further_mfr/p3.1A_extract_smoking_followup_for_model.R (parallelisation over task ID 1 - 100).

p2_data_prep_further_mfr/p3.2A_extract_prescriptions_followup_for_model.R (parallelisation over task ID 1 - 100, and statin/antihypertensive).

p4_validate/p1_prototype3_calculate_cf_surv_times.R (parallelisation over task ID 1 – 200, and male/female).

p4_validate/p4.1_instability_fit_models.R (parallelisation over task ID 1 – 500, and male/female).

### 3. Choice of predictors in the model

Three changes were made from the predictors included in QRISK4, which is the basis for the predictors included in the initial risk estimation layer. Non-HDL cholesterol was chosen rather than total cholesterol/HDL ratio because this is a variable which we plan to intervene on, but there is a lack of evidence in the literature about the effect of intervening on total cholesterol/HDL ratio on cardiovascular risk. Furthermore, there is a lack of evidence on the effect of our interventions (such as statins and antihypertensives) on total cholesterol/HDL ratio. There was evidence around the effect of non-HDL cholesterol on cardiovascular risk, and its effects with respect to the other modifiable risk factors, and therefore we chose to include this. For the same reasons, systolic blood pressure variability was excluded as a predictor. This is a variable we would have been intervening on, but there was a lack of evidence about the effect reducing systolic blood pressure variability on cardiovascular risk. It would therefore not possible to include these in the intervention layer and the DAG. Finally, calendar time was included as a predictor due to the secular trend in cardiovascular disease observed in the data (see supplementary data file 2). When implementing the model in practice, we would recommend making all predicts at the maximum follow-up time date, which was 01/03/2020. The numeric value for this is 5537 (number of days from 01/01/2005).

### 4. Methodology for model evaluation with respect to the imputation procedure

The imputation procedure results in 10 development datasets and therefore 10 different risk prediction models. There are also 10 validation datasets. Under normal circumstances, in each validation dataset, the risks from the 10 models could be pooled, resulting in one of each performance metrics in each validation dataset. However, when assessing calibration and discrimination, counterfactual survival times which align with the treatment regime of interest (no change to intervention status during follow-up) must be estimated. The process for doing so is dependent on the cumulative baseline hazard of the model being evaluated. Is it unclear how to pool the cumulative baseline hazards of the different developed models, and the impact this may have on the estimation of the counterfactual survival times. Furthermore, for a given development dataset and resulting model, the counterfactual survival times vary across each of the validation datasets given the imputation of the smoking status variable at baseline, which impacts what is deemed a change in treatment.

We therefore followed recommendations for when measures cannot be pooled^18^ and report imputation-specific performance metrics. This results in a different performance metrics (whether calibration plots, ICI, E50, E90 or C-statistic) for every pair of development and validation dataset, resulting in 100 performance metrics. We follow further recommendations^19^ and report robust summary measures (median, and 2.5^th^ and 97.5^th^ percentiles) of the model performance metrics across these 100 imputed development/validation dataset pairs. We initially did this for the model evaluation in the entire validation cohort, but found minimal difference in performance across each pair (see Figure 3 from the manuscript, or section 4.3.1 in supplementary data file 2). Therefore all other evaluation analyses (assessment of fairness, temporal validation) were done only in one development and validation pair.

In Figure 3 from the manuscript, the analysis of observed vs predicted within 100 groups defined by predicted risk, is also the median value for each group across every combination of development and validation dataset.

### 5. Definition of time-varying variables used to adjust for treatment drop-in

We use an approach inspired by Xu et al.^20^ In the referenced study, they create a time-varying variable which is split after an individual initiates statin treatment, and fix the coefficient of this variable to be the effect of initiating statins. In this study, we have a scenario where individuals may be on or off treatment at baseline, and may move on and off multiple treatments more than once. Importantly, the Estimand of this model is the risk of staying on the current intervention strategy (with respect to statins, antihypertensives and an individual’s smoking status), rather than the risk under the treatment strategy of not receiving the intervention. The process to augment the process of Xu et al., into this setting is as follows.

Let $t_{i}, i\in\{1,2,\ldots,n\}$ be all the times at which an individual changes status with respect to statin use, antihypertensive use or smoking status. Let $t_{event}$ be the time at which an individual either has a cardiovascular outcome event or is censored.

We define four variables. Statin use at baseline ($A_{stat}=0/1$). Antihypertensive use at baseline ($A_{ah}=0/1$). Smoking initiation status at baseline ($A_{smokinit}=0$ if never smoker, 1 if ex or current smoker). Smoking cessation status at baseline ($A_{smokcess}=0$ if never smoker or current smoker, 1 if ex-smoker). For individuals with missing smoking status at baseline, we use the imputed value.

We then define four-time varying variables, which is the individuals treatment status during follow-up. Statin use at time $t$ ($A_{stat}(t)=0/1$). Antihypertensive use at time $t$ ($A_{ah}(t)=0/1$). Smoking initiation status at time $t$ ($A_{smokinit}(t)=0$ if never smoker, 1 if ex or current smoker). Smoking cessation status at time $t$ ($A_{smokcess}(t)=0$ if never smoker or current smoker, 1 if ex-smoker).

We then define four new time varying variables, which is the individual’s treatment status during follow-up, relative to their baseline value.

$${AR}_{stat}\left( t \right)=A_{stat}\left( t \right)-A_{stat}$$

$${AR}_{ah}\left( t \right)=A_{ah}\left( t \right)-A_{ah}$$

$${AR}_{smokinit}\left( t \right)=A_{smokinit}\left( t \right)-A_{smokinit}$$

$${AR}_{smokcess}\left( t \right)=A_{smokcess}\left( t \right)-A_{smokcess}$$

Finally, we define new time-varying variables, which is the intervention status relative to their value at baseline, multiplied by the log of the total effect of that intervention ($B$), in the form of a log-hazard ratio.

$$B_{stat}\left( t \right)={AR}_{stat}\left( t \right)*B_{stat}$$

$$B_{ah}\left( t \right)={AR}_{ah}\left( t \right)*B_{ah}$$

$$B_{smokinit}\left( t \right)={AR}_{smokinit}\left( t \right)*B_{smokinit}$$

$$B_{smokcess}\left( t \right)={AR}_{smokcess}\left( t \right)*B_{smokcess}$$

Where $B_{stat}=-0.3102413$, $B_{ah}=-0.3245535$, $B_{smokinit}=0.3410332$, $B_{smokcess}=-0.2080456$.

### 6. Methodology for estimation of counterfactual survival times

We again use an approach inspired by Xu et al.^20^ In the referenced study, they obtain treatment naive counter factual survival times by adjusting the cumulative baseline hazard dependent after an individual initiates statin treatment, to get the cumulative hazard that would have been observed if the individual had not initiated statins. A new counterfactual survival times is then estimated based off this augmented cumulative baseline hazard.

In this study, we have a scenario where individuals may be on or off treatment at baseline, and may move on and off multiple treatments more than once. Importantly, the Estimand of this model is the risk of staying on the current intervention strategy (with respect to statins, antihypertensives and an individual’s smoking status), rather than the risk under the treatment strategy of not receiving the intervention. The process to augment the process of Xu et al., into this setting is as follows.

Firstly, the time-varying variables are calculated as detailed in section 5. The cumulative hazard is then calculated at each time point $t_{i}$: $H_{0}\left( t_{i} \right)$.

The counterfactual cumulative hazard at time $t$ is then estimated as:

$$H_{cf}\left( t \right)=H\left( t_{1} \right)+\sum_{i=1}^{n} \exp\left( B_{stat}\left( t_{i} \right)+B_{ah}\left( t_{i} \right)+B_{smokinit}\left( t_{i} \right)+B_{smokcess}\left( t_{i} \right) \right)*H(t_{i+1})-\exp\left( B_{stat}\left( t_{i} \right)+B_{ah}\left( t_{i} \right)+B_{smokinit}\left( t_{i} \right)+B_{smokcess}\left( t_{i} \right) \right)*H(t_{i})$$

Where $t_{n+1}=t_{event}$ is the time at individual is either censored or has a cardiovascular outcome event.

In Plain English Terms, the cumulative hazard is broken up into intervals, defined by when an individual changes treatment status. If the treatment status during an interval is different from what it was at zero, the amount of cumulative hazard in that interval is adjusted to the amount of hazard if the treatment status had been the same as at baseline. The values of time-varying variables at the times $t_{i}$, $B_{stat}\left( t_{i} \right)$, $B_{ah}\left( t_{i} \right)$, $B_{smokinit}\left( t_{i} \right)$ and $B_{smokcess}\left( t_{i} \right)$, are the value they take for the upcoming interval $[t_{i},t_{i+1})$.

The counterfactual survival time, $t_{cf}$, is then estimated in the same way as Xu et al.,^20^ by finding the minimum time point at which $H\left( t \right)=H_{cf}\left( t_{event} \right)$. For individuals where $t_{cf}$ is bigger than the maximum follow-up, we set $t_{cf}$ to be the value at maximum follow-up. The event indicator remains the same.

It should also be noted that this process is dependent on the cumulative hazard function, which changes across each of the models fitted in the different multiply imputed development datasets. Normally, for model evaluation it is suggested to average the predicted survival probabilities from each of the models using Rubins Rules, and assess the performance of these ‘averaged’ risks in each of the validation datasets. However, given the need to estimate counterfactual survival times is dependent on the cumulative hazard, it is unclear how to do this with the ‘averaged risks’. We therefore initially estimated performance in each pair of development/validation datasets. After finding minimal differences in performance across these pairs, for all fairness checks and temporal validation, these were just done in 1 development/validation pair.

### 8. CHARIOT architecture with N visits

This diagram has been included to highlight that at visit N, the change in the modifiable risk factors is always calculated relative to the values recorded at visit 0.

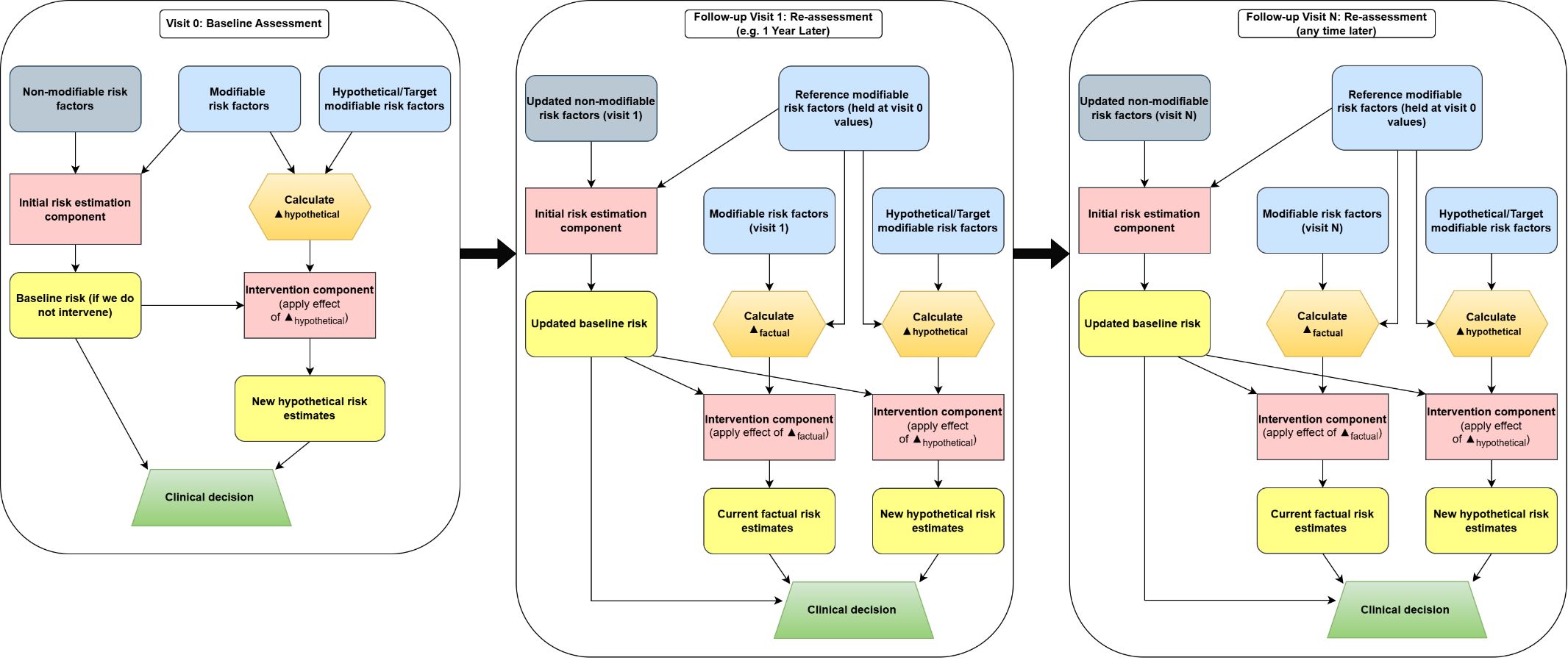

9.	Hippisley-Cox J, Coupland C, Brindle P. Development and validation of QRISK3 risk prediction algorithms to estimate future risk of cardiovascular disease: prospective cohort study. *BMJ*; 357.

10.	QResearch. QResearch: QCode Group Library, https://www.qresearch.org/data/qcode-group-library/ (2024, accessed 12 February 2024).

11.	Head A. GitHub: Annalhead/CPRD_multimorbidity_codelists, https://github.com/annalhead/CPRD_multimorbidity_codelists (accessed 26 November 2021).

12.	Head A, Fleming K, Kypridemos C, et al. Inequalities in incident and prevalent multimorbidity in England, 2004–19: a population-based, descriptive study. *Lancet Heal Longev* 2021; 2: e489–e497.

13.	Muzambi R, Bhaskaran K, Strongman H, et al. Trends and inequalities in statin use for the primary and secondary prevention of cardiovascular disease between 2009 and 2021 in England Authors: Rutendo Muzambi. *medRxiv*. Epub ahead of print 2024. DOI: 10.1101/2024.11.22.24317782.

14.	Greater Manchester Integrated Digital Care Record. Greater Manchester Integrated Digital Care Record: Clinical Code Sets, https://github.com/rw251/gm-idcr/tree/master/shared/clinical-code-sets/conditions.
