## Supplementary data files 1, 2 and 3 for "Predicting cardiovascular risk under intervention: Development and internal validation of the CHARIOT Model in 19 million adults": CHARIOT applied - Supplementary data file 2 - results.html


### Supplementary data file 2 - results

#### 2025-05-14

- 1 Baseline information
  - 1.1 Female cohort
  - 1.2 Male cohort
  - 1.3 Overall outcome incidence
    rates
- 2 Convergence plots for
  imputation
- 3 Shape of non-linear effects
  interacted with age
  - 3.1 Interaction with binary
    variables
  - 3.2 Interaction with polytomous
    variables
  - 3.3 Interaction with continuous
    variables
  - 3.4 Hazard ratio for age
- 4 Model evaluation at baseline (visit
  0)
  - 4.1 Calibration plots
    - 4.1.1 Calibration in the entire
      cohort
    - 4.1.2 Calibration by age
    - 4.1.3 Calibration by
      ethnicity
    - 4.1.4 Calibration by region
  - 4.2 Calibration tables
  - 4.3 Discrimination
    - 4.3.1 Entire cohort
    - 4.3.2 Across subgroups
  - 4.4 Instability plots
- 5 Temporal validation of initial risk
  estimation layer
  - 5.1 Calibration plots
    - 5.1.1 Female
    - 5.1.2 Male
  - 5.2 Calibration tables
  - 5.3 Discrimination
- 6 Temporal validation of intervention
  layer
  - 6.1 Calibration plots
    - 6.1.1 SBP
    - 6.1.2 BMI
    - 6.1.3 Non-HDL cholesterol
    - 6.1.4 Smoking status
  - 6.2 Calibration tables
  - 6.3 Discrimination

### 1 Baseline information

#### 1.1 Female cohort

Baseline information

Characteristics at baseline index date

| **Characteristic** | **development**  N = 6,929,696^1^ | **validation**  N = 2,969,869^1^ |
| --- | --- | --- |
| gender |  |  |
| 2 | 6,929,696 (100%) | 2,969,869 (100%) |
| age |  |  |
| Median (5% Centile, Q1, Q3, 95% Centile) | 34 (18, 25, 49, 73) | 34 (18, 25, 49, 73) |
| N Missing (% Missing%) | 0 (0%) | 0 (0%) |
| ethnicity |  |  |
| bangladeshi | 43,277 (0.6%) | 18,378 (0.6%) |
| black african | 181,931 (2.6%) | 78,495 (2.6%) |
| black caribbean | 85,845 (1.2%) | 36,784 (1.2%) |
| chinese | 114,966 (1.7%) | 49,099 (1.7%) |
| indian | 184,284 (2.7%) | 78,619 (2.6%) |
| other asian | 151,978 (2.2%) | 65,218 (2.2%) |
| other ethnic | 339,518 (4.9%) | 146,198 (4.9%) |
| pakistani | 98,819 (1.4%) | 42,487 (1.4%) |
| white | 4,910,925 (71%) | 2,104,181 (71%) |
| Missing | 818,153 (12%) | 350,410 (12%) |
| hypertension | 590,168 (8.5%) | 253,150 (8.5%) |
| ra | 41,340 (0.6%) | 17,859 (0.6%) |
| af | 31,866 (0.5%) | 13,636 (0.5%) |
| ckd | 48,832 (0.7%) | 20,942 (0.7%) |
| smi | 106,151 (1.5%) | 45,653 (1.5%) |
| fhcvd | 489,757 (7.1%) | 209,389 (7.1%) |
| migraine | 491,209 (7.1%) | 210,680 (7.1%) |
| sle | 8,378 (0.1%) | 3,473 (0.1%) |
| diabetes |  |  |
| Absent | 6,764,724 (98%) | 2,898,508 (98%) |
| Type1 | 22,967 (0.3%) | 10,099 (0.3%) |
| Type2 | 142,005 (2.0%) | 61,262 (2.1%) |
| impotence | 1,325 (<0.1%) | 568 (<0.1%) |
| cortico | 50,139 (0.7%) | 21,508 (0.7%) |
| antipsy | 26,998 (0.4%) | 11,768 (0.4%) |
| smoking |  |  |
| Non-smoker | 3,700,998 (53%) | 1,586,524 (53%) |
| Ex-smoker | 1,350,555 (19%) | 577,256 (19%) |
| Current | 1,303,909 (19%) | 560,242 (19%) |
| Missing | 574,234 (8.3%) | 245,847 (8.3%) |
| statins | 223,438 (3.2%) | 96,352 (3.2%) |
| antihypertensives | 553,177 (8.0%) | 237,712 (8.0%) |
| copd | 55,568 (0.8%) | 23,891 (0.8%) |
| int\_dis | 21,499 (0.3%) | 9,343 (0.3%) |
| downs | 3,470 (<0.1%) | 1,396 (<0.1%) |
| oral\_cancer | 2,285 (<0.1%) | 1,035 (<0.1%) |
| brain\_cancer | 1,818 (<0.1%) | 785 (<0.1%) |
| lung\_cancer | 2,706 (<0.1%) | 1,038 (<0.1%) |
| blood\_cancer | 13,681 (0.2%) | 5,863 (0.2%) |
| pre\_eclampsia | 19,431 (0.3%) | 8,358 (0.3%) |
| postnatal\_depression | 91,692 (1.3%) | 39,140 (1.3%) |
| bmi |  |  |
| Median (5% Centile, Q1, Q3, 95% Centile) | 24.2 (19.1, 21.6, 28.3, 36.6) | 24.2 (19.1, 21.6, 28.3, 36.6) |
| N Missing (% Missing%) | 2,507,999 (36%) | 1,075,506 (36%) |
| sbp |  |  |
| Median (5% Centile, Q1, Q3, 95% Centile) | 120 (99, 110, 131, 150) | 120 (99, 110, 131, 150) |
| N Missing (% Missing%) | 1,430,104 (21%) | 612,157 (21%) |
| sbp\_var |  |  |
| Median (5% Centile, Q1, Q3, 95% Centile) | 8.7 (1.0, 5.7, 12.7, 19.9) | 8.7 (1.0, 5.6, 12.7, 19.9) |
| N Missing (% Missing%) | 3,191,936 (46%) | 1,367,343 (46%) |
| cholhdl\_ratio |  |  |
| Median (5% Centile, Q1, Q3, 95% Centile) | 3.40 (2.10, 2.80, 4.20, 5.60) | 3.40 (2.10, 2.80, 4.20, 5.60) |
| N Missing (% Missing%) | 5,738,652 (83%) | 2,460,544 (83%) |
| nonhdl |  |  |
| Median (5% Centile, Q1, Q3, 95% Centile) | 3.60 (2.10, 2.90, 4.40, 5.70) | 3.60 (2.10, 2.90, 4.40, 5.70) |
| N Missing (% Missing%) | 5,745,669 (83%) | 2,463,590 (83%) |
| IMD |  |  |
| Median (5% Centile, Q1, Q3, 95% Centile) | 11.0 (2.0, 6.0, 15.0, 19.0) | 11.0 (2.0, 6.0, 15.0, 19.0) |
| N Missing (% Missing%) | 8,544 (0.1%) | 3,741 (0.1%) |
| cholesterol |  |  |
| Median (5% Centile, Q1, Q3, 95% Centile) | 5.20 (3.60, 4.50, 6.00, 7.20) | 5.20 (3.60, 4.50, 6.00, 7.20) |
| N Missing (% Missing%) | 5,505,444 (79%) | 2,360,651 (79%) |
| hdl |  |  |
| Median (5% Centile, Q1, Q3, 95% Centile) | 1.50 (0.99, 1.26, 1.80, 2.38) | 1.50 (0.99, 1.26, 1.80, 2.38) |
| N Missing (% Missing%) | 5,748,636 (83%) | 2,464,770 (83%) |
| ldl |  |  |
| Median (5% Centile, Q1, Q3, 95% Centile) | 3.00 (1.66, 2.40, 3.70, 4.80) | 3.00 (1.67, 2.40, 3.70, 4.80) |
| N Missing (% Missing%) | 5,985,980 (86%) | 2,566,826 (86%) |
|  |  |  |
| --- | --- | --- |
| ^1^ n (%) | | |

Outcome incidence rates

| total follow up (years) | n events | rate (per 1000 years) | year |
| --- | --- | --- | --- |
| 3549114 | 19530 | 5.502781 | 2005-01-01 |
| 3623014 | 19314 | 5.330920 | 2006-01-01 |
| 3693905 | 19232 | 5.206414 | 2007-01-01 |
| 3787201 | 19668 | 5.193282 | 2008-01-01 |
| 3855245 | 19658 | 5.099027 | 2009-01-01 |
| 3914935 | 19596 | 5.005447 | 2010-01-01 |
| 3969437 | 19485 | 4.908757 | 2011-01-01 |
| 4045228 | 20074 | 4.962390 | 2012-01-01 |
| 4079327 | 19421 | 4.760835 | 2013-01-01 |
| 4111786 | 19433 | 4.726170 | 2014-01-01 |
| 4198558 | 19709 | 4.694231 | 2015-01-01 |
| 4321645 | 19841 | 4.591076 | 2016-01-01 |
| 4418138 | 20555 | 4.652412 | 2017-01-01 |
| 4513019 | 20766 | 4.601354 | 2018-01-01 |
| 5256346 | 25101 | 4.775371 | 2019-01-01 |

#### 1.2 Male cohort

Baseline information

Characteristics at baseline index date

| **Characteristic** | **development**  N = 6,657,587^1^ | **validation**  N = 2,853,251^1^ |
| --- | --- | --- |
| gender |  |  |
| 1 | 6,657,587 (100%) | 2,853,251 (100%) |
| age |  |  |
| Median (5% Centile, Q1, Q3, 95% Centile) | 35 (18, 25, 48, 69) | 35 (18, 25, 48, 69) |
| N Missing (% Missing%) | 0 (0%) | 0 (0%) |
| ethnicity |  |  |
| bangladeshi | 47,739 (0.7%) | 20,343 (0.7%) |
| black african | 161,076 (2.4%) | 68,622 (2.4%) |
| black caribbean | 69,313 (1.0%) | 29,520 (1.0%) |
| chinese | 76,534 (1.1%) | 33,081 (1.2%) |
| indian | 186,961 (2.8%) | 80,134 (2.8%) |
| other asian | 148,388 (2.2%) | 63,788 (2.2%) |
| other ethnic | 297,217 (4.5%) | 127,462 (4.5%) |
| pakistani | 109,090 (1.6%) | 46,543 (1.6%) |
| white | 4,161,907 (63%) | 1,783,483 (63%) |
| Missing | 1,399,362 (21%) | 600,275 (21%) |
| hypertension | 491,513 (7.4%) | 210,718 (7.4%) |
| ra | 15,948 (0.2%) | 6,729 (0.2%) |
| af | 42,997 (0.6%) | 18,482 (0.6%) |
| ckd | 34,740 (0.5%) | 14,888 (0.5%) |
| smi | 92,766 (1.4%) | 39,917 (1.4%) |
| fhcvd | 373,041 (5.6%) | 159,804 (5.6%) |
| migraine | 189,737 (2.8%) | 81,834 (2.9%) |
| sle | 1,059 (<0.1%) | 461 (<0.1%) |
| diabetes |  |  |
| Absent | 6,457,605 (97%) | 2,767,670 (97%) |
| Type1 | 30,724 (0.5%) | 12,986 (0.5%) |
| Type2 | 169,258 (2.5%) | 72,595 (2.5%) |
| impotence | 264,625 (4.0%) | 113,546 (4.0%) |
| cortico | 30,086 (0.5%) | 12,980 (0.5%) |
| antipsy | 33,183 (0.5%) | 14,445 (0.5%) |
| smoking |  |  |
| Non-smoker | 2,808,997 (42%) | 1,204,336 (42%) |
| Ex-smoker | 1,218,152 (18%) | 522,006 (18%) |
| Current | 1,533,440 (23%) | 657,095 (23%) |
| Missing | 1,096,998 (16%) | 469,814 (16%) |
| statins | 236,194 (3.5%) | 100,852 (3.5%) |
| antihypertensives | 439,853 (6.6%) | 188,251 (6.6%) |
| copd | 58,969 (0.9%) | 25,101 (0.9%) |
| int\_dis | 31,114 (0.5%) | 13,533 (0.5%) |
| downs | 3,644 (<0.1%) | 1,543 (<0.1%) |
| oral\_cancer | 4,030 (<0.1%) | 1,702 (<0.1%) |
| brain\_cancer | 2,165 (<0.1%) | 941 (<0.1%) |
| lung\_cancer | 2,852 (<0.1%) | 1,194 (<0.1%) |
| blood\_cancer | 16,121 (0.2%) | 6,822 (0.2%) |
| pre\_eclampsia | 14 (<0.1%) | 6 (<0.1%) |
| postnatal\_depression | 230 (<0.1%) | 104 (<0.1%) |
| bmi |  |  |
| Median (5% Centile, Q1, Q3, 95% Centile) | 25.4 (19.8, 22.8, 28.5, 34.5) | 25.4 (19.8, 22.8, 28.5, 34.5) |
| N Missing (% Missing%) | 3,133,025 (47%) | 1,341,453 (47%) |
| sbp |  |  |
| Median (5% Centile, Q1, Q3, 95% Centile) | 130 (105, 120, 139, 155) | 130 (105, 120, 139, 155) |
| N Missing (% Missing%) | 2,781,539 (42%) | 1,191,013 (42%) |
| sbp\_var |  |  |
| Median (5% Centile, Q1, Q3, 95% Centile) | 9 (1, 5, 14, 21) | 9 (0, 5, 14, 21) |
| N Missing (% Missing%) | 4,920,835 (74%) | 2,109,056 (74%) |
| cholhdl\_ratio |  |  |
| Median (5% Centile, Q1, Q3, 95% Centile) | 4.04 (2.40, 3.29, 5.00, 6.50) | 4.03 (2.40, 3.29, 4.96, 6.50) |
| N Missing (% Missing%) | 5,511,264 (83%) | 2,361,129 (83%) |
| nonhdl |  |  |
| Median (5% Centile, Q1, Q3, 95% Centile) | 3.79 (2.11, 3.05, 4.50, 5.70) | 3.78 (2.11, 3.05, 4.50, 5.70) |
| N Missing (% Missing%) | 5,516,764 (83%) | 2,363,442 (83%) |
| IMD |  |  |
| Median (5% Centile, Q1, Q3, 95% Centile) | 11.0 (2.0, 6.0, 15.0, 19.0) | 11.0 (2.0, 6.0, 15.0, 19.0) |
| N Missing (% Missing%) | 8,251 (0.1%) | 3,582 (0.1%) |
| cholesterol |  |  |
| Median (5% Centile, Q1, Q3, 95% Centile) | 5.10 (3.40, 4.40, 5.80, 7.00) | 5.10 (3.40, 4.40, 5.80, 7.00) |
| N Missing (% Missing%) | 5,296,726 (80%) | 2,269,252 (80%) |
| hdl |  |  |
| Median (5% Centile, Q1, Q3, 95% Centile) | 1.22 (0.80, 1.04, 1.50, 1.95) | 1.22 (0.80, 1.04, 1.50, 1.95) |
| N Missing (% Missing%) | 5,519,844 (83%) | 2,364,826 (83%) |
| ldl |  |  |
| Median (5% Centile, Q1, Q3, 95% Centile) | 3.07 (1.60, 2.40, 3.70, 4.70) | 3.06 (1.60, 2.40, 3.70, 4.70) |
| N Missing (% Missing%) | 5,756,144 (86%) | 2,466,236 (86%) |
|  |  |  |
| --- | --- | --- |
| ^1^ n (%) | | |

Outcome incidence rates

| total follow up (years) | n events | rate (per 1000 years) | year |
| --- | --- | --- | --- |
| 3554268 | 24999 | 7.033517 | 2005-01-01 |
| 3619813 | 25113 | 6.937652 | 2006-01-01 |
| 3683994 | 24809 | 6.734267 | 2007-01-01 |
| 3762988 | 24937 | 6.626914 | 2008-01-01 |
| 3814630 | 25500 | 6.684789 | 2009-01-01 |
| 3863668 | 25204 | 6.523335 | 2010-01-01 |
| 3897504 | 25269 | 6.483380 | 2011-01-01 |
| 3962134 | 25887 | 6.533600 | 2012-01-01 |
| 3975906 | 25474 | 6.407093 | 2013-01-01 |
| 4023383 | 25565 | 6.354105 | 2014-01-01 |
| 4107558 | 26005 | 6.331012 | 2015-01-01 |
| 4228502 | 26767 | 6.330137 | 2016-01-01 |
| 4333320 | 27809 | 6.417481 | 2017-01-01 |
| 4437280 | 28048 | 6.320990 | 2018-01-01 |
| 5180255 | 33674 | 6.500452 | 2019-01-01 |

#### 1.3 Overall outcome incidence rates

Overall:

| Gender | total follow up (years) | n events | rate (per 1000 years) |
| --- | --- | --- | --- |
| Male | 60445203 | 395060 | 6.535837 |
| Female | 61336898 | 301383 | 4.913568 |

Primary care only:

| Gender | total follow up (years) | n events | rate (per 1000 years) |
| --- | --- | --- | --- |
| Male | 60665210 | 299952 | 4.944382 |
| Female | 61540711 | 221315 | 3.596237 |

Secondary care only:

| Gender | total follow up (years) | n events | rate (per 1000 years) |
| --- | --- | --- | --- |
| Male | 60898361 | 269188 | 4.420283 |
| Female | 61786335 | 187618 | 3.036561 |

ONS death data only:

| Gender | total follow up (years) | n events | rate (per 1000 years) |
| --- | --- | --- | --- |
| Male | 62211142 | 66633 | 1.0710782 |
| Female | 62597055 | 52057 | 0.8316206 |

### 2 Convergence plots for imputation

Female

Male

### 3 Shape of non-linear effects interacted with age

Hazard ratio as age increases, interacted with each of the other
predictor variables. The spline for age had knots at 25, 40, 55 and
70.

#### 3.1 Interaction with binary variables

#### 3.2 Interaction with polytomous variables

*Female Hazard Ratios*

*Male Hazard Ratios*

#### 3.3 Interaction with continuous variables

These plots show how the hazrd ratio of the variable of interst
changes relative to a reference value, for a discrete range of ages. The
reference value is 25 for BMI, 120 for SBP, 5 for SBP variability, and 3
for cholesterol/HDL ratio. Plotting from the 2.5th to p7.5th
percentile.

Alternatively, I could plot how the hazard ratio comparing two fixed
values (say 40 vs 25, 30 vs 25, and 20 vs 25) changes as age increases.
This would arguably be more ‘similar’ to the other plots, but I think
less informative.

*Female Hazard Ratios*

*Male Hazard Ratios*

#### 3.4 Hazard ratio for age

Hazard ratio is assessed at the mean of all other continuous
variables, and the reference value for all categorical variables. the
hazard ratio is relative to someone age 40.

### 4 Model evaluation at baseline (visit 0)

#### 4.1 Calibration plots

##### 4.1.1 Calibration in the entire cohort

Female:

Male:

##### 4.1.2 Calibration by age

Female:

Male:

##### 4.1.3 Calibration by ethnicity

Female:

Male:

##### 4.1.4 Calibration by region

Female:

Male:

#### 4.2 Calibration tables

Female:

|  | N | ICI | E50 | E90 |
| --- | --- | --- | --- | --- |
| Entire cohort | 2969869 | 0.001 | 0.000 | 0.003 |
| White | 2383595 | 0.001 | 0.000 | 0.003 |
| Bangladeshi | 20838 | 0.003 | 0.001 | 0.005 |
| Black african | 89036 | 0.001 | 0.001 | 0.002 |
| Black caribbean | 41817 | 0.002 | 0.001 | 0.006 |
| Chinese | 56055 | 0.000 | 0.000 | 0.001 |
| Indian | 89690 | 0.001 | 0.000 | 0.004 |
| Other asian | 74230 | 0.001 | 0.001 | 0.001 |
| Other ethnic | 166417 | 0.001 | 0.000 | 0.000 |
| Pakistani | 48191 | 0.001 | 0.001 | 0.002 |
| (17,30] | 1212938 | 0.000 | 0.000 | 0.001 |
| (30,40] | 636504 | 0.001 | 0.000 | 0.001 |
| (40,50] | 406103 | 0.001 | 0.000 | 0.003 |
| (50,60] | 305530 | 0.002 | 0.001 | 0.003 |
| (60,70] | 213948 | 0.005 | 0.004 | 0.008 |
| (70,80] | 137794 | 0.009 | 0.009 | 0.011 |
| (80,Inf] | 57052 | 0.007 | 0.005 | 0.015 |
| East Midlands | 84623 | 0.001 | 0.000 | 0.006 |
| East of England | 121992 | 0.001 | 0.001 | 0.003 |
| London | 708877 | 0.001 | 0.000 | 0.001 |
| North East | 83788 | 0.002 | 0.001 | 0.007 |
| North West | 485399 | 0.005 | 0.001 | 0.017 |
| South Central | 352873 | 0.002 | 0.001 | 0.005 |
| South East Coast | 244682 | 0.001 | 0.000 | 0.001 |
| South West | 344630 | 0.001 | 0.000 | 0.002 |
| West Midlands | 428709 | 0.002 | 0.001 | 0.003 |
| Yorkshire And The Humber | 111098 | 0.003 | 0.000 | 0.010 |

Male:

|  | N | ICI | E50 | E90 |
| --- | --- | --- | --- | --- |
| Entire cohort | 2969869 | 0.001 | 0.000 | 0.003 |
| White | 2383595 | 0.001 | 0.000 | 0.003 |
| Bangladeshi | 20838 | 0.003 | 0.001 | 0.005 |
| Black african | 89036 | 0.001 | 0.001 | 0.002 |
| Black caribbean | 41817 | 0.002 | 0.001 | 0.006 |
| Chinese | 56055 | 0.000 | 0.000 | 0.001 |
| Indian | 89690 | 0.001 | 0.000 | 0.004 |
| Other asian | 74230 | 0.001 | 0.001 | 0.001 |
| Other ethnic | 166417 | 0.001 | 0.000 | 0.000 |
| Pakistani | 48191 | 0.001 | 0.001 | 0.002 |
| (17,30] | 1212938 | 0.000 | 0.000 | 0.001 |
| (30,40] | 636504 | 0.001 | 0.000 | 0.001 |
| (40,50] | 406103 | 0.001 | 0.000 | 0.003 |
| (50,60] | 305530 | 0.002 | 0.001 | 0.003 |
| (60,70] | 213948 | 0.005 | 0.004 | 0.008 |
| (70,80] | 137794 | 0.009 | 0.009 | 0.011 |
| (80,Inf] | 57052 | 0.007 | 0.005 | 0.015 |
| East Midlands | 84623 | 0.001 | 0.000 | 0.006 |
| East of England | 121992 | 0.001 | 0.001 | 0.003 |
| London | 708877 | 0.001 | 0.000 | 0.001 |
| North East | 83788 | 0.002 | 0.001 | 0.007 |
| North West | 485399 | 0.005 | 0.001 | 0.017 |
| South Central | 352873 | 0.002 | 0.001 | 0.005 |
| South East Coast | 244682 | 0.001 | 0.000 | 0.001 |
| South West | 344630 | 0.001 | 0.000 | 0.002 |
| West Midlands | 428709 | 0.002 | 0.001 | 0.003 |
| Yorkshire And The Humber | 111098 | 0.003 | 0.000 | 0.010 |

#### 4.3 Discrimination

##### 4.3.1 Entire cohort

The mean (sd) and median (2.5th - 97.5th percentile range) across the
results when validated in each development/validation pair.

Female:

| mean (sd) | median (p2.5, p97.5) | min | max |
| --- | --- | --- | --- |
| 0.87 (0) | 0.87 (0.87,0.87) | 0.87 | 0.87 |

Male:

| mean (sd) | median (p2.5, p97.5) | min | max |
| --- | --- | --- | --- |
| 0.86 (0) | 0.86 (0.86,0.86) | 0.86 | 0.86 |

##### 4.3.2 Across subgroups

Evaluated in one development/validation pair.

Female:

|  | N | C\_index | index\_lower | index\_upper |
| --- | --- | --- | --- | --- |
| Entire cohort | 2969869 | 0.874 | 1.08795e+11 | 1.244425e+11 |
| white | 2383595 | 0.868 | 8.68000e-01 | 8.680000e-01 |
| bangladeshi | 20838 | 0.902 | 9.01000e-01 | 9.020000e-01 |
| black african | 89036 | 0.871 | 8.71000e-01 | 8.710000e-01 |
| black caribbean | 41817 | 0.858 | 8.57000e-01 | 8.580000e-01 |
| chinese | 56055 | 0.918 | 9.17000e-01 | 9.180000e-01 |
| indian | 89690 | 0.894 | 8.94000e-01 | 8.950000e-01 |
| other asian | 74230 | 0.903 | 9.03000e-01 | 9.030000e-01 |
| other ethnic | 166417 | 0.893 | 8.93000e-01 | 8.930000e-01 |
| pakistani | 48191 | 0.894 | 8.94000e-01 | 8.940000e-01 |
| (17,30] | 1212938 | 0.643 | 6.43000e-01 | 6.430000e-01 |
| (30,40] | 636504 | 0.738 | 7.38000e-01 | 7.380000e-01 |
| (40,50] | 406103 | 0.731 | 7.31000e-01 | 7.310000e-01 |
| (50,60] | 305530 | 0.681 | 6.81000e-01 | 6.810000e-01 |
| (60,70] | 213948 | 0.637 | 6.37000e-01 | 6.370000e-01 |
| (70,80] | 137794 | 0.603 | 6.03000e-01 | 6.030000e-01 |
| (80,Inf] | 57052 | 0.577 | 5.77000e-01 | 5.770000e-01 |
| East Midlands | 84623 | 0.886 | 8.85000e-01 | 8.860000e-01 |
| East of England | 121992 | 0.872 | 8.72000e-01 | 8.720000e-01 |
| London | 708877 | 0.893 | 8.93000e-01 | 8.930000e-01 |
| North East | 83788 | 0.864 | 8.64000e-01 | 8.640000e-01 |
| North West | 485399 | 0.864 | 8.64000e-01 | 8.640000e-01 |
| South Central | 352873 | 0.882 | 8.82000e-01 | 8.820000e-01 |
| South East Coast | 244682 | 0.873 | 8.73000e-01 | 8.730000e-01 |
| South West | 344630 | 0.868 | 8.68000e-01 | 8.680000e-01 |
| West Midlands | 428709 | 0.859 | 8.59000e-01 | 8.590000e-01 |
| Yorkshire And The Humber | 111098 | 0.876 | 8.76000e-01 | 8.760000e-01 |

Male:

|  | N | C\_index | index\_lower | index\_upper |
| --- | --- | --- | --- | --- |
| Entire cohort | 2853251 | 0.859 | 1.39642e+11 | 1.6261e+11 |
| white | 2255357 | 0.850 | 8.50000e-01 | 8.5000e-01 |
| bangladeshi | 25778 | 0.893 | 8.93000e-01 | 8.9300e-01 |
| black african | 87189 | 0.875 | 8.75000e-01 | 8.7600e-01 |
| black caribbean | 37750 | 0.882 | 8.82000e-01 | 8.8200e-01 |
| chinese | 42249 | 0.937 | 9.37000e-01 | 9.3700e-01 |
| indian | 101988 | 0.886 | 8.86000e-01 | 8.8600e-01 |
| other asian | 81291 | 0.892 | 8.92000e-01 | 8.9200e-01 |
| other ethnic | 162327 | 0.879 | 8.79000e-01 | 8.7900e-01 |
| pakistani | 59322 | 0.893 | 8.93000e-01 | 8.9300e-01 |
| (17,30] | 1085269 | 0.702 | 7.02000e-01 | 7.0200e-01 |
| (30,40] | 667332 | 0.722 | 7.22000e-01 | 7.2200e-01 |
| (40,50] | 466242 | 0.693 | 6.93000e-01 | 6.9300e-01 |
| (50,60] | 313025 | 0.650 | 6.50000e-01 | 6.5000e-01 |
| (60,70] | 192902 | 0.610 | 6.10000e-01 | 6.1100e-01 |
| (70,80] | 99173 | 0.587 | 5.87000e-01 | 5.8700e-01 |
| (80,Inf] | 29308 | 0.568 | 5.68000e-01 | 5.6800e-01 |
| East Midlands | 82136 | 0.880 | 8.80000e-01 | 8.8000e-01 |
| East of England | 115336 | 0.851 | 8.51000e-01 | 8.5100e-01 |
| London | 669396 | 0.882 | 8.82000e-01 | 8.8200e-01 |
| North East | 80533 | 0.842 | 8.42000e-01 | 8.4200e-01 |
| North West | 474969 | 0.850 | 8.50000e-01 | 8.5000e-01 |
| South Central | 338673 | 0.859 | 8.59000e-01 | 8.5900e-01 |
| South East Coast | 232467 | 0.850 | 8.50000e-01 | 8.5000e-01 |
| South West | 329698 | 0.854 | 8.54000e-01 | 8.5400e-01 |
| West Midlands | 418996 | 0.845 | 8.45000e-01 | 8.4500e-01 |
| Yorkshire And The Humber | 107727 | 0.859 | 8.58000e-01 | 8.5900e-01 |

#### 4.4 Instability plots

These instability pare plotted for 3,000 randomly selected
individuals. The variation on the y-axis represents the expected
variability in risk if the development cohort was to be re-sampled from
the entire population which it represents, and a model was developed in
these re-sampled cohorts. Naturally, variation increases as the
predicted risk increases. We see a small number of individuals where the
variability exceeds that of other individuals at their level of
predicted risk. Looking at these individuals highlights them to be those
with a higher number of risk factors.

### 5 Temporal validation of initial risk estimation layer

Follow-up index dates are defined at 1/2/3/4/5 years after baseline.
Calibration and discrimination are assessed at 5 and 10 years.

In plots and tables, t\_fup = number of days after baseline the index
date is defined at, t\_eval is denoting whether evaluating 5 or 10 year
risk predictions.

#### 5.1 Calibration plots

##### 5.1.1 Female

##### 5.1.2 Male

#### 5.2 Calibration tables

| gender | t | follow-up visit | N | ICI | E50 | E90 |
| --- | --- | --- | --- | --- | --- | --- |
| female | 5 | 1 | 2455459 | 0.000 | 0.000 | 0.000 |
| female | 5 | 2 | 2058812 | 0.001 | 0.000 | 0.003 |
| female | 5 | 3 | 1757932 | 0.002 | 0.001 | 0.004 |
| female | 5 | 4 | 1532580 | 0.002 | 0.001 | 0.006 |
| female | 5 | 5 | 1351932 | 0.003 | 0.001 | 0.007 |
| female | 10 | 1 | 2455459 | 0.001 | 0.000 | 0.002 |
| female | 10 | 2 | 2058812 | 0.003 | 0.001 | 0.007 |
| female | 10 | 3 | 1757932 | 0.003 | 0.001 | 0.009 |
| female | 10 | 4 | 1532580 | 0.004 | 0.002 | 0.011 |
| female | 10 | 5 | 1351932 | 0.005 | 0.002 | 0.013 |
| male | 5 | 1 | 2425374 | 0.001 | 0.000 | 0.001 |
| male | 5 | 2 | 2077642 | 0.001 | 0.001 | 0.003 |
| male | 5 | 3 | 1793566 | 0.002 | 0.001 | 0.004 |
| male | 5 | 4 | 1562777 | 0.002 | 0.001 | 0.004 |
| male | 5 | 5 | 1374424 | 0.002 | 0.001 | 0.004 |
| male | 10 | 1 | 2425374 | 0.001 | 0.001 | 0.003 |
| male | 10 | 2 | 2077642 | 0.003 | 0.002 | 0.006 |
| male | 10 | 3 | 1793566 | 0.004 | 0.002 | 0.008 |
| male | 10 | 4 | 1562777 | 0.004 | 0.003 | 0.009 |
| male | 10 | 5 | 1374424 | 0.005 | 0.003 | 0.010 |

#### 5.3 Discrimination

| gender | t | follow-up visit | N | C\_index | index\_lower | index\_upper |
| --- | --- | --- | --- | --- | --- | --- |
| female | 5 years | 1 | 2455459 | 0.867 | 0.867 | 0.867 |
| female | 5 years | 2 | 2058812 | 0.859 | 0.859 | 0.859 |
| female | 5 years | 3 | 1757932 | 0.853 | 0.853 | 0.853 |
| female | 5 years | 4 | 1532580 | 0.847 | 0.847 | 0.847 |
| female | 5 years | 5 | 1351932 | 0.841 | 0.841 | 0.841 |
| female | 10 years | 1 | 2455459 | 0.867 | 0.867 | 0.867 |
| female | 10 years | 2 | 2058812 | 0.859 | 0.859 | 0.859 |
| female | 10 years | 3 | 1757932 | 0.853 | 0.853 | 0.853 |
| female | 10 years | 4 | 1532580 | 0.847 | 0.847 | 0.847 |
| female | 10 years | 5 | 1351932 | 0.841 | 0.841 | 0.841 |
| male | 5 years | 1 | 2425374 | 0.850 | 0.850 | 0.850 |
| male | 5 years | 2 | 2077642 | 0.842 | 0.842 | 0.842 |
| male | 5 years | 3 | 1793566 | 0.834 | 0.834 | 0.834 |
| male | 5 years | 4 | 1562777 | 0.826 | 0.826 | 0.826 |
| male | 5 years | 5 | 1374424 | 0.818 | 0.818 | 0.818 |
| male | 10 years | 1 | 2425374 | 0.850 | 0.850 | 0.850 |
| male | 10 years | 2 | 2077642 | 0.842 | 0.842 | 0.842 |
| male | 10 years | 3 | 1793566 | 0.834 | 0.834 | 0.834 |
| male | 10 years | 4 | 1562777 | 0.826 | 0.826 | 0.826 |
| male | 10 years | 5 | 1374424 | 0.818 | 0.818 | 0.818 |

### 6 Temporal validation of intervention layer

Follow-up index dates are defined at 1/2/3/4/5 years after baseline.
Calibration and discrimination are assessed at 5 and 10 years. We assess
interventional changes in each modifiable risk factor sequentially.

In plots and tables, t\_fup = number of days after baseline the index
date is defined at, t\_eval is denoting whether evaluating 5 or 10 year
risk predictions, mfr denotes the modifiable risk factor being
evaluated.

#### 6.1 Calibration plots

##### 6.1.1 SBP

###### 6.1.1.1 Female

###### 6.1.1.2 Male

##### 6.1.2 BMI

###### 6.1.2.1 Female

###### 6.1.2.2 Male

##### 6.1.3 Non-HDL cholesterol

###### 6.1.3.1 Female

###### 6.1.3.2 Male

##### 6.1.4 Smoking status

###### 6.1.4.1 Female

###### 6.1.4.2 Male

#### 6.2 Calibration tables

| gender | t | MFR | follow-up visit | N | ICI | E50 | E90 |
| --- | --- | --- | --- | --- | --- | --- | --- |
| female | 5 | systolic blood pressure | 1 | 1968593 | 0.002 | 0.000 | 0.005 |
| female | 5 | systolic blood pressure | 2 | 1662971 | 0.003 | 0.000 | 0.004 |
| female | 5 | systolic blood pressure | 3 | 1428062 | 0.003 | 0.000 | 0.004 |
| female | 5 | systolic blood pressure | 4 | 1250995 | 0.004 | 0.000 | 0.004 |
| female | 5 | systolic blood pressure | 5 | 1105753 | 0.004 | 0.001 | 0.006 |
| female | 5 | body mass index | 1 | 1543950 | 0.001 | 0.000 | 0.001 |
| female | 5 | body mass index | 2 | 1269513 | 0.001 | 0.000 | 0.003 |
| female | 5 | body mass index | 3 | 1062159 | 0.002 | 0.000 | 0.004 |
| female | 5 | body mass index | 4 | 909644 | 0.002 | 0.001 | 0.006 |
| female | 5 | body mass index | 5 | 788406 | 0.002 | 0.001 | 0.007 |
| female | 5 | non-HDL cholesterol | 1 | 428588 | 0.004 | 0.004 | 0.005 |
| female | 5 | non-HDL cholesterol | 2 | 366484 | 0.004 | 0.003 | 0.006 |
| female | 5 | non-HDL cholesterol | 3 | 316919 | 0.004 | 0.003 | 0.008 |
| female | 5 | non-HDL cholesterol | 4 | 276939 | 0.004 | 0.003 | 0.009 |
| female | 5 | non-HDL cholesterol | 5 | 241240 | 0.004 | 0.003 | 0.009 |
| female | 5 | smoking status | 1 | 2255786 | 0.000 | 0.000 | 0.000 |
| female | 5 | smoking status | 2 | 1891789 | 0.001 | 0.000 | 0.003 |
| female | 5 | smoking status | 3 | 1614590 | 0.002 | 0.001 | 0.005 |
| female | 5 | smoking status | 4 | 1406987 | 0.003 | 0.001 | 0.007 |
| female | 5 | smoking status | 5 | 1239519 | 0.003 | 0.001 | 0.008 |
| female | 10 | systolic blood pressure | 1 | 1968593 | 0.004 | 0.000 | 0.008 |
| female | 10 | systolic blood pressure | 2 | 1662971 | 0.005 | 0.001 | 0.007 |
| female | 10 | systolic blood pressure | 3 | 1428062 | 0.006 | 0.001 | 0.007 |
| female | 10 | systolic blood pressure | 4 | 1250995 | 0.006 | 0.001 | 0.010 |
| female | 10 | systolic blood pressure | 5 | 1105753 | 0.006 | 0.001 | 0.013 |
| female | 10 | body mass index | 1 | 1543950 | 0.001 | 0.000 | 0.002 |
| female | 10 | body mass index | 2 | 1269513 | 0.002 | 0.001 | 0.006 |
| female | 10 | body mass index | 3 | 1062159 | 0.003 | 0.001 | 0.009 |
| female | 10 | body mass index | 4 | 909644 | 0.004 | 0.001 | 0.011 |
| female | 10 | body mass index | 5 | 788406 | 0.005 | 0.001 | 0.014 |
| female | 10 | non-HDL cholesterol | 1 | 428588 | 0.007 | 0.007 | 0.010 |
| female | 10 | non-HDL cholesterol | 2 | 366484 | 0.008 | 0.006 | 0.015 |
| female | 10 | non-HDL cholesterol | 3 | 316919 | 0.008 | 0.006 | 0.019 |
| female | 10 | non-HDL cholesterol | 4 | 276939 | 0.008 | 0.005 | 0.019 |
| female | 10 | non-HDL cholesterol | 5 | 241240 | 0.008 | 0.005 | 0.021 |
| female | 10 | smoking status | 1 | 2255786 | 0.001 | 0.000 | 0.001 |
| female | 10 | smoking status | 2 | 1891789 | 0.003 | 0.001 | 0.007 |
| female | 10 | smoking status | 3 | 1614590 | 0.004 | 0.001 | 0.011 |
| female | 10 | smoking status | 4 | 1406987 | 0.005 | 0.002 | 0.013 |
| female | 10 | smoking status | 5 | 1239519 | 0.006 | 0.002 | 0.016 |
| male | 5 | systolic blood pressure | 1 | 1416229 | 0.004 | 0.001 | 0.008 |
| male | 5 | systolic blood pressure | 2 | 1215758 | 0.004 | 0.001 | 0.008 |
| male | 5 | systolic blood pressure | 3 | 1052251 | 0.005 | 0.001 | 0.008 |
| male | 5 | systolic blood pressure | 4 | 919573 | 0.005 | 0.001 | 0.007 |
| male | 5 | systolic blood pressure | 5 | 808802 | 0.006 | 0.002 | 0.007 |
| male | 5 | body mass index | 1 | 1260371 | 0.001 | 0.001 | 0.003 |
| male | 5 | body mass index | 2 | 1053639 | 0.001 | 0.001 | 0.001 |
| male | 5 | body mass index | 3 | 885211 | 0.002 | 0.001 | 0.003 |
| male | 5 | body mass index | 4 | 751650 | 0.002 | 0.001 | 0.004 |
| male | 5 | body mass index | 5 | 644397 | 0.002 | 0.001 | 0.004 |
| male | 5 | non-HDL cholesterol | 1 | 415115 | 0.004 | 0.005 | 0.007 |
| male | 5 | non-HDL cholesterol | 2 | 354341 | 0.004 | 0.004 | 0.005 |
| male | 5 | non-HDL cholesterol | 3 | 304912 | 0.004 | 0.004 | 0.006 |
| male | 5 | non-HDL cholesterol | 4 | 264177 | 0.004 | 0.004 | 0.006 |
| male | 5 | non-HDL cholesterol | 5 | 228701 | 0.005 | 0.004 | 0.007 |
| male | 5 | smoking status | 1 | 2021227 | 0.001 | 0.000 | 0.002 |
| male | 5 | smoking status | 2 | 1724289 | 0.001 | 0.000 | 0.002 |
| male | 5 | smoking status | 3 | 1481106 | 0.002 | 0.001 | 0.003 |
| male | 5 | smoking status | 4 | 1283919 | 0.002 | 0.001 | 0.004 |
| male | 5 | smoking status | 5 | 1122313 | 0.002 | 0.001 | 0.005 |
| male | 10 | systolic blood pressure | 1 | 1416229 | 0.006 | 0.001 | 0.012 |
| male | 10 | systolic blood pressure | 2 | 1215758 | 0.007 | 0.001 | 0.011 |
| male | 10 | systolic blood pressure | 3 | 1052251 | 0.008 | 0.002 | 0.012 |
| male | 10 | systolic blood pressure | 4 | 919573 | 0.008 | 0.002 | 0.015 |
| male | 10 | systolic blood pressure | 5 | 808802 | 0.009 | 0.002 | 0.018 |
| male | 10 | body mass index | 1 | 1260371 | 0.002 | 0.001 | 0.004 |
| male | 10 | body mass index | 2 | 1053639 | 0.003 | 0.001 | 0.004 |
| male | 10 | body mass index | 3 | 885211 | 0.004 | 0.002 | 0.008 |
| male | 10 | body mass index | 4 | 751650 | 0.004 | 0.002 | 0.011 |
| male | 10 | body mass index | 5 | 644397 | 0.005 | 0.002 | 0.011 |
| male | 10 | non-HDL cholesterol | 1 | 415115 | 0.007 | 0.008 | 0.011 |
| male | 10 | non-HDL cholesterol | 2 | 354341 | 0.007 | 0.007 | 0.011 |
| male | 10 | non-HDL cholesterol | 3 | 304912 | 0.008 | 0.006 | 0.016 |
| male | 10 | non-HDL cholesterol | 4 | 264177 | 0.008 | 0.006 | 0.018 |
| male | 10 | non-HDL cholesterol | 5 | 228701 | 0.009 | 0.006 | 0.020 |
| male | 10 | smoking status | 1 | 2021227 | 0.001 | 0.001 | 0.002 |
| male | 10 | smoking status | 2 | 1724289 | 0.002 | 0.001 | 0.005 |
| male | 10 | smoking status | 3 | 1481106 | 0.004 | 0.002 | 0.009 |
| male | 10 | smoking status | 4 | 1283919 | 0.005 | 0.003 | 0.011 |
| male | 10 | smoking status | 5 | 1122313 | 0.005 | 0.003 | 0.013 |

#### 6.3 Discrimination

| gender | t | MFR | follow-up visit | N | C\_index | index\_lower | index\_upper |
| --- | --- | --- | --- | --- | --- | --- | --- |
| female | 5 years | systolic blood pressure | 1 | 1968593 | 0.857 | 0.857 | 0.857 |
| female | 5 years | systolic blood pressure | 2 | 1662971 | 0.849 | 0.849 | 0.849 |
| female | 5 years | systolic blood pressure | 3 | 1428062 | 0.842 | 0.842 | 0.842 |
| female | 5 years | systolic blood pressure | 4 | 1250995 | 0.837 | 0.837 | 0.837 |
| female | 5 years | systolic blood pressure | 5 | 1105753 | 0.831 | 0.831 | 0.831 |
| female | 5 years | body mass index | 1 | 1543950 | 0.868 | 0.868 | 0.868 |
| female | 5 years | body mass index | 2 | 1269513 | 0.861 | 0.861 | 0.861 |
| female | 5 years | body mass index | 3 | 1062159 | 0.855 | 0.855 | 0.855 |
| female | 5 years | body mass index | 4 | 909644 | 0.849 | 0.849 | 0.849 |
| female | 5 years | body mass index | 5 | 788406 | 0.844 | 0.844 | 0.844 |
| female | 5 years | non-HDL cholesterol | 1 | 428588 | 0.757 | 0.757 | 0.757 |
| female | 5 years | non-HDL cholesterol | 2 | 366484 | 0.751 | 0.751 | 0.751 |
| female | 5 years | non-HDL cholesterol | 3 | 316919 | 0.746 | 0.746 | 0.746 |
| female | 5 years | non-HDL cholesterol | 4 | 276939 | 0.743 | 0.743 | 0.743 |
| female | 5 years | non-HDL cholesterol | 5 | 241240 | 0.739 | 0.738 | 0.739 |
| female | 5 years | smoking status | 1 | 2255786 | 0.865 | 0.865 | 0.865 |
| female | 5 years | smoking status | 2 | 1891789 | 0.857 | 0.857 | 0.857 |
| female | 5 years | smoking status | 3 | 1614590 | 0.851 | 0.851 | 0.851 |
| female | 5 years | smoking status | 4 | 1406987 | 0.845 | 0.845 | 0.845 |
| female | 5 years | smoking status | 5 | 1239519 | 0.839 | 0.839 | 0.839 |
| female | 10 years | systolic blood pressure | 1 | 1968593 | 0.858 | 0.858 | 0.858 |
| female | 10 years | systolic blood pressure | 2 | 1662971 | 0.850 | 0.850 | 0.850 |
| female | 10 years | systolic blood pressure | 3 | 1428062 | 0.843 | 0.843 | 0.843 |
| female | 10 years | systolic blood pressure | 4 | 1250995 | 0.838 | 0.838 | 0.838 |
| female | 10 years | systolic blood pressure | 5 | 1105753 | 0.832 | 0.832 | 0.832 |
| female | 10 years | body mass index | 1 | 1543950 | 0.868 | 0.868 | 0.868 |
| female | 10 years | body mass index | 2 | 1269513 | 0.861 | 0.861 | 0.861 |
| female | 10 years | body mass index | 3 | 1062159 | 0.855 | 0.855 | 0.855 |
| female | 10 years | body mass index | 4 | 909644 | 0.849 | 0.849 | 0.849 |
| female | 10 years | body mass index | 5 | 788406 | 0.844 | 0.844 | 0.844 |
| female | 10 years | non-HDL cholesterol | 1 | 428588 | 0.757 | 0.757 | 0.757 |
| female | 10 years | non-HDL cholesterol | 2 | 366484 | 0.751 | 0.751 | 0.751 |
| female | 10 years | non-HDL cholesterol | 3 | 316919 | 0.746 | 0.746 | 0.746 |
| female | 10 years | non-HDL cholesterol | 4 | 276939 | 0.743 | 0.743 | 0.743 |
| female | 10 years | non-HDL cholesterol | 5 | 241240 | 0.739 | 0.739 | 0.739 |
| female | 10 years | smoking status | 1 | 2255786 | 0.865 | 0.865 | 0.865 |
| female | 10 years | smoking status | 2 | 1891789 | 0.857 | 0.857 | 0.857 |
| female | 10 years | smoking status | 3 | 1614590 | 0.851 | 0.851 | 0.851 |
| female | 10 years | smoking status | 4 | 1406987 | 0.845 | 0.845 | 0.845 |
| female | 10 years | smoking status | 5 | 1239519 | 0.839 | 0.839 | 0.839 |
| male | 5 years | systolic blood pressure | 1 | 1416229 | 0.816 | 0.816 | 0.816 |
| male | 5 years | systolic blood pressure | 2 | 1215758 | 0.806 | 0.806 | 0.806 |
| male | 5 years | systolic blood pressure | 3 | 1052251 | 0.797 | 0.797 | 0.797 |
| male | 5 years | systolic blood pressure | 4 | 919573 | 0.788 | 0.788 | 0.788 |
| male | 5 years | systolic blood pressure | 5 | 808802 | 0.779 | 0.779 | 0.779 |
| male | 5 years | body mass index | 1 | 1260371 | 0.844 | 0.844 | 0.844 |
| male | 5 years | body mass index | 2 | 1053639 | 0.834 | 0.834 | 0.834 |
| male | 5 years | body mass index | 3 | 885211 | 0.825 | 0.825 | 0.825 |
| male | 5 years | body mass index | 4 | 751650 | 0.817 | 0.817 | 0.817 |
| male | 5 years | body mass index | 5 | 644397 | 0.808 | 0.808 | 0.808 |
| male | 5 years | non-HDL cholesterol | 1 | 415115 | 0.736 | 0.736 | 0.736 |
| male | 5 years | non-HDL cholesterol | 2 | 354341 | 0.729 | 0.729 | 0.729 |
| male | 5 years | non-HDL cholesterol | 3 | 304912 | 0.722 | 0.722 | 0.722 |
| male | 5 years | non-HDL cholesterol | 4 | 264177 | 0.716 | 0.716 | 0.716 |
| male | 5 years | non-HDL cholesterol | 5 | 228701 | 0.708 | 0.708 | 0.708 |
| male | 5 years | smoking status | 1 | 2021227 | 0.843 | 0.843 | 0.843 |
| male | 5 years | smoking status | 2 | 1724289 | 0.833 | 0.833 | 0.833 |
| male | 5 years | smoking status | 3 | 1481106 | 0.824 | 0.824 | 0.824 |
| male | 5 years | smoking status | 4 | 1283919 | 0.816 | 0.816 | 0.816 |
| male | 5 years | smoking status | 5 | 1122313 | 0.807 | 0.807 | 0.807 |
| male | 10 years | systolic blood pressure | 1 | 1416229 | 0.816 | 0.816 | 0.816 |
| male | 10 years | systolic blood pressure | 2 | 1215758 | 0.807 | 0.807 | 0.807 |
| male | 10 years | systolic blood pressure | 3 | 1052251 | 0.798 | 0.798 | 0.798 |
| male | 10 years | systolic blood pressure | 4 | 919573 | 0.789 | 0.789 | 0.789 |
| male | 10 years | systolic blood pressure | 5 | 808802 | 0.781 | 0.781 | 0.781 |
| male | 10 years | body mass index | 1 | 1260371 | 0.844 | 0.844 | 0.844 |
| male | 10 years | body mass index | 2 | 1053639 | 0.834 | 0.834 | 0.834 |
| male | 10 years | body mass index | 3 | 885211 | 0.825 | 0.825 | 0.825 |
| male | 10 years | body mass index | 4 | 751650 | 0.817 | 0.817 | 0.817 |
| male | 10 years | body mass index | 5 | 644397 | 0.808 | 0.808 | 0.808 |
| male | 10 years | non-HDL cholesterol | 1 | 415115 | 0.736 | 0.736 | 0.736 |
| male | 10 years | non-HDL cholesterol | 2 | 354341 | 0.729 | 0.729 | 0.729 |
| male | 10 years | non-HDL cholesterol | 3 | 304912 | 0.722 | 0.722 | 0.722 |
| male | 10 years | non-HDL cholesterol | 4 | 264177 | 0.716 | 0.716 | 0.716 |
| male | 10 years | non-HDL cholesterol | 5 | 228701 | 0.709 | 0.709 | 0.709 |
| male | 10 years | smoking status | 1 | 2021227 | 0.843 | 0.843 | 0.843 |
| male | 10 years | smoking status | 2 | 1724289 | 0.833 | 0.833 | 0.833 |
| male | 10 years | smoking status | 3 | 1481106 | 0.824 | 0.824 | 0.824 |
| male | 10 years | smoking status | 4 | 1283919 | 0.816 | 0.816 | 0.816 |
| male | 10 years | smoking status | 5 | 1122313 | 0.807 | 0.807 | 0.807 |
